# *NTRK* oncogenic fusions are exclusively associated with the serrated neoplasia pathway in the colorectum and begin to occur in sessile serrated lesions

**DOI:** 10.1101/2021.03.29.21253871

**Authors:** Jung Ho Kim, Jeong Hoon Hong, Yoon-La Choi, Ji Ae Lee, Mi-kyoung Seo, Mi-Sook Lee, Sung Bin An, Min Jung Sung, Nam-Yun Cho, Sung-Su Kim, Young Kee Shin, Sangwoo Kim, Gyeong Hoon Kang

**Affiliations:** Department of Pathology, Seoul National University Hospital, Seoul National University College of Medicine, Seoul, Korea; Laboratory of Epigenetics, Cancer Research Institute, Seoul National University College of Medicine, Seoul, Korea; Central Laboratory, LOGONE Bio-Convergence Research Foundation, Seoul, Korea; Department of Health Sciences and Technology, Samsung Advanced Institute for Health Sciences & Technology, Sungkyunkwan University, Seoul, Korea; Laboratory of Cancer Genomics and Molecular Pathology, Samsung Medical Center, Sungkyunkwan University School of Medicine, Seoul, Korea; Department of Pathology and Translational Genomics, Samsung Medical Center, Sungkyunkwan University School of Medicine, Seoul, Korea; Department of Biomedical Systems Informatics, Yonsei University College of Medicine, Seoul, Korea; Brain Korea 21 PLUS Project for Medical Science, Yonsei University College of Medicine, Seoul, Korea; Department of Molecular Medicine and Biopharmaceutical Sciences, Graduate School of Convergence Science and Technology, Seoul National University, Seoul, Korea

**Keywords:** colorectal cancer, colorectal adenomas, colonic polyps, molecular carcinogenesis, colorectal pathology

## Abstract

**Background:** *NTRK* fusions are emerging tissue-agnostic drug targets in malignancies including colorectal cancers (CRCs), but their detailed landscape in the context of various colorectal carcinogenesis pathways remains to be investigated.

**Methods:** Pan-TRK expression was assessed by immunohistochemistry in retrospectively collected colorectal epithelial tumor tissues, including 441 CRCs (133 microsatellite instability-high (MSI-high) and 308 microsatellite stable (MSS)) and 565 premalignant colorectal lesions (300 serrated lesions and 265 conventional adenomas). TRK-positive cases were subjected to next-generation sequencing and/or fluorescence in situ hybridization to confirm *NTRK* rearrangements.

**Results:** TRK positivity was not observed in any of the MSS CRCs, conventional adenomas, traditional serrated adenomas, or hyperplastic polyps, whereas TRK positivity was observed in 11 of 58 (19%) sporadic MSI-high CRCs, 4 of 23 (17%) sessile serrated lesions with dysplasia (SSLDs), and 5 of 132 (4%) SSLs. The 11 TRK-positive MSI-high CRCs commonly harbored CpG island methylator phenotype-high (CIMP-high), *MLH1* methylation, *KRAS*/*BRAF* wild-type, and *NTRK1* or *NTRK3* fusion (*TPM3*-*NTRK1, TPR*-*NTRK1, LMNA*-*NTRK1, SFPQ*-*NTRK1, ETV6*-*NTRK3*, or *EML4*-*NTRK3*). Both *NTRK1* or *NTRK3* rearrangement and *KRAS*/*BRAF* wild-type were detected in all nine TRK-positive SSL(D)s, seven of which demonstrated MSS and/or CIMP-low. TRK overexpression and early dysplastic changes are occasionally co-localized in the crypt base area of SSLs. Age-related occurrence patterns suggest that the progression interval from *NTRK*-rearranged SSLs to CRCs may be shorter than from *BRAF*-mutated SSLs to CRCs.

**Conclusion:** *NTRK*-rearranged colorectal tumors develop exclusively through the serrated neoplasia pathway and can be initiated from non-dysplastic SSLs without *KRAS*/*BRAF* mutations prior to full occurrence of MSI-high/CIMP-high.

## INTRODUCTION

Tropomyosin receptor kinase (TRK) proteins belong to one of the receptor tyrosine kinase families, including TRKA, TRKB, and TRKC, encoded by neurotrophic tropomyosin receptor kinase 1 (*NTRK1*), *NTRK2*, and *NTRK3*, respectively.^1^ Although the normal balanced function of TRK receptors is physiologically important particularly in the development of the nervous system,^2, 3^ constitutive activation of TRK receptors contributes to carcinogenesis because constitutive activation of their downstream signals, including the RAS/MAPK/ERK and PI3K/AKT pathways, can lead to an uncontrolled increase in cellular proliferation and survival.^4, 5^ In fact, aberrant activation of TRK receptors by *NTRK* gene fusions is known to be a major oncogenic driver in subsets of human malignancies.^1^ More importantly, in recent years, *NTRK* fusions have emerged as druggable targets in various tumors, and TRK inhibitors, including larotrectinib and entrectinib, have been recently approved by the U.S. Food and Drug Administration for the treatment of adult and pediatric patients with *NTRK* fusion-positive cancers regardless of tumor type.^6, 7^ *NTRK* fusions are the second approved histology-agnostic biomarker for cancer treatment following the first histology-agnostic approval of microsatellite instability (MSI) as a predictive biomarker for cancer immunotherapy using immune checkpoint inhibitors.^6^

Although *NTRK* fusions in colorectal cancers (CRCs) are rare (< 1%),^6-10^ characteristic molecular associations of *NTRK* fusion-positive CRCs have been identified in recent studies using next-generation sequencing (NGS). In detail, major oncogenic gene fusions, including *NTRK* fusions, have been found specifically in a sporadic *MLH1*-methylated, MSI-high, *KRAS*/*BRAF* wild-type subset among CRCs.^11-16^ These findings imply that *NTRK* fusions selectively occur in CRCs developed from serrated premalignant lesions because sessile serrated lesions (SSLs) are recognized as major precursors of sporadic MSI-high CRCs.^17-20^ However, until recently, there have been no reports revealing the detailed association between *NTRK* fusions and specific colorectal carcinogenesis pathways, including the serrated neoplasia pathway and the conventional neoplasia pathway.

Therefore, we decided to investigate the comprehensive clinicopathological and molecular landscape of TRK expression and *NTRK* fusions in a large series of colorectal premalignant lesions as well as CRCs. Because it has been acknowledged that pan-TRK immunohistochemistry (IHC) can be a reliable tool for the screening of *NTRK* fusions in various tumor tissues,^21-24^ we utilized pan-TRK IHC in 565 premalignant lesions and 441 CRCs as an initial step of this study. Next, TRK-positive tumors identified by pan-TRK IHC were subjected to next-generation sequencing (NGS) or fluorescence in situ hybridization (FISH) to identify *NTRK* gene fusions or translocations.

## MATERIALS AND METHODS

### Tissue samples

All tissue samples were retrospectively collected from the pathology archives of Seoul National University Hospital (SNUH) in Seoul, Korea. Detailed methods of sample collection are summarized in Supplementary Figure S1. A consecutive series of 133 primary CRCs with microsatellite instability-high (MSI-high) were retrieved from archived formalin-fixed, paraffin-embedded (FFPE) and fresh frozen (FF) (if available) specimens that had been surgically resected between 2014 and 2018 at SNUH. Another consecutive series of 308 primary microsatellite stable (MSS) CRCs, which had been surgically resected in 2018 at SNUH, were also selected. A total of 565 premalignant colorectal lesions, including 132 SSLs, 23 SSLs with dysplasia (SSLDs), 45 traditional serrated adenomas (TSAs), 100 hyperplastic polyps (HPs), 150 conventional adenomas with low-grade dysplasia (CALGs), and 115 conventional adenomas with high-grade dysplasia (CAHGs), were retrieved from archived FFPE tissues that had been endoscopically resected between 2010 and 2012 or surgically resected between 2016 and 2018 at SNUH. Histological classification of the 565 premalignant lesions was conducted through microscopic review of hematoxylin and eosin (H&E)-stained tissue slides of all cases by experienced gastrointestinal pathologists (J.H.K., J.A.L., and G.H.K.). The three pathologists independently assessed histological subtypes of the premalignant lesions using criteria from the latest World Health Organization (WHO) classification of the digestive system (5^th^ edition),^25^ and the final diagnosis of each case was reached by agreement between the three pathologists. The 132 SSLs without morphological dysplasia were collected in our previous study, and the detailed histological criteria for the selection of SSLs were described in our previous reports.^18, 19^ Other precursor lesions, including SSLDs, TSAs, HPs, CALGs, and CAHGs, were selected mainly based on the histological criteria of the latest WHO classification.^19, 20, 25, 26^ This study was conducted in compliance with the ethical guidelines of the 2013 Declaration of Helsinki. The Institutional Review Board (IRB) of SNUH approved this study (IRB No. 1804-036-935; 1804-109-939). All CRCs and some premalignant lesions included in this study, which were surgically resected between 2014 and 2018, were previously reposited in the Cancer Tissue Bank of SNUH with informed consent obtained from all patients. For premalignant lesions endoscopically resected between 2010 and 2012, obtaining informed consent from patients was waived by the IRB because this was a retrospective, anonymized, tissue-based investigation. The case number used in this study (MCRC-X, SSLD-X, or SSL-X) is a sample number anonymized for research only and does not display any identifiable information about the patient.

### Clinicopathological data

Clinical data, including age, sex, and tumor location, of the 441 CRCs, 132 SSLs, and 23 SSLDs were collected through a review of electronic medical records. The histopathological features of CRCs, including tumor differentiation (grade), histological components (mucinous, signet ring cell, or medullary), lymphatic/venous/perineural invasion, tumor budding, poorly differentiated clusters, desmoplastic reaction, and tertiary lymphoid structures, were reviewed by two gastrointestinal pathologists (J.H.K. and J.A.L.). Quantification of the densities of tumor-infiltrating immune cells, including CD3^+^ T cells, CD8^+^ T cells, FoxP3^+^ T cells, CD68^+^ macrophages, and CD163^+^ macrophages, in CRCs was conducted using the QuPath-based computational image analysis method as previously described.^27^

### Immunohistochemistry

To efficiently perform IHC, tissue microarray (TMA) blocks of the 133 MSI-high CRCs, 308 MSS CRCs, 132 SSLs, 23 SSLDs, 45 TSAs, 100 HPs, 150 CALGs, and 115 CAHGs were constructed as previously described.^27^ IHC using two different antibody clones against pan-TRK (EPR17341 clone, Abcam, Cambridge, UK; and A7H6R clone, Cell Signaling Technology, Danvers, MA, USA) was conducted on TMA slides of all of the 441 CRCs and 565 premalignant lesions. IHC staining using anti-MLH1 antibody (M1 clone, Ventana RTU, Roche, Basel, Switzerland) or anti-ALK antibody (D5F3 CDx Assay, Ventana RTU, Roche) was also performed on TMA slides of the 441 CRCs, 132 SSLs, and 23 SSLDs. Pan-TRK expression positivity in a case was determined as nuclear and/or cytoplasmic staining in at least 5% of tumor or lesion epithelial cells with any of the 3-tier intensities (1+/2+/3+), and a case showing positivity in at least one stain among the two pan-TRK IHC stains using EPR17341 or A7H6R clone antibodies was finally included in TRK-positive cases in the present study. Loss of MLH1 expression was determined as negative staining in the nuclei of tumor or lesion epithelial cells, in contrast to internal positive controls such as nuclear staining in normal epithelial cells or immune cells.

### Microsatellite instability analysis

All 441 CRCs, 132 SSLs, and 23 SSLDs were subjected to capillary electrophoresis DNA fragment analysis-based MSI testing using the Bethesda guideline-recommended microsatellite markers (BAT-25, BAT-26, D5S346, D17S250, and D2S123).^28^ MSI status of each sample was determined as MSI-high or MSS (including MSI-low). The MSI-high phenotype was diagnosed when two or more microsatellite markers of tumor DNA showed instability compared with normal mucosa DNA in a case.

### DNA methylation analysis

Genomic DNA was isolated from formalin-fixed, paraffin-embedded (FFPE) tissues of the 133 MSI-high CRCs, 308 MSS CRCs, 132 SSLs, and 23 SSLDs. Subsequent bisulfite modification of the DNA samples was conducted as previously described.^17^ CpG island methylator phenotype (CIMP) status of each CRC, SSL, or SSLD was determined by the methylation-specific real-time PCR method (MethyLight assay) using eight CIMP-specific gene promoter markers (*MLH1, NEUROG1, CRABP1, CACNA1G, CDKN2A* (*p16*), *IGF2, RUNX3*, and *SOCS1*).^17^ If a case demonstrated CpG island hypermethylation in five or more markers, the case was classified as CIMP-high. Cases with none of the hypermethylated markers were determined to be CIMP-negative. The remaining cases (one to four hypermethylated markers) were categorized as CIMP-low.

### *KRAS*/*BRAF* mutation analysis

The mutation status of *KRAS* exons 2, 3, and 4 and *BRAF* exon 15 in the 441 CRCs, 132 SSLs, and 23 SSLDs were analysed by Sanger sequencing as previously described.^17^

### Targeted RNA sequencing

For targeted RNA sequencing of 12 samples, RNA was isolated from FFPE and/or fresh frozen (FF) tissues of nine TRK-positive CRCs (MCRC-4, 12, 27, 48, 55, 83, 113, 120, and 133) and three TRK-positive SSL(D)s (SSLD-4, 10, and SSL-10) using the RNeasy FFPE Kit (QIAGEN, Hilden, Germany) according to the manufacturer’s instructions. After isolation of total RNA, ribosomal RNA (rRNA) was depleted with RNase H, using the NEBNext® rRNA Depletion kit (NEW ENGLAND BioLabs Inc., MA, USA) according to the manufacturer’s protocols. RNA without rRNA was measured for concentration and purity using a Qubit Fluorometer (Thermo Fisher Scientific, MA, USA) and 2200 TapeStation System (Agilent, CA, USA), respectively. *NTRK* fusions were detected using a SOLIDaccuTest™ RNA Kit (NGeneBio Inc., Seoul, Korea), a commercialized fusion partner-agnostic NGS panel kit. Briefly, after the prepared RNA was converted to cDNA, sequencing libraries were constructed along a series of steps for adaptor ligation, size selection, and PCR enrichment, according to the manufacturer’s protocol. Target enrichment based on hybridization capture was performed using the SOLIDaccuTest™ RNA Kit, which targets CDS regions of *NTRK1, NTRK2*, and *NTRK3*. Library quality and quantity were measured using the Qubit Fluorometer and 2200 TapeStation System. Additional PCR enrichment was performed, and paired-end sequencing (2 × 150 bp) was performed using a MiSeq Dx sequencer (Illumina, CA, USA) according to the manufacturer’s instructions. All procedures of the targeted RNA sequencing were conducted in the central laboratory of the LOGONE Bio-Convergence Research Foundation, Seoul, Korea.

### Whole transcriptome and exome sequencing

For whole transcriptome sequencing of three samples (MCRC-27, 43, and 71), 100 ng total RNA was isolated from paired tumor and normal FF tissues of three TRK-positive CRCs and was subjected to sequencing library preparation using the TruSeq stranded total RNA sample preparation kit (Illumina, CA, USA) which combines RiboZero rRNA depletion with a stranded-specific method similar to the dUDP method. The quality of these cDNA libraries was evaluated using an Agilent 2100 Bioanalyzer (Agilent, CA, USA). They were quantified using the KAPA library quantification kit (Kapa Biosystems, MA, USA) according to the manufacturer’s library quantification protocol. Following cluster amplification of denatured templates, sequencing was performed as paired-end (2 × 100 bp) using a NovaSeq 6000 sequencer (Illumina, CA, USA).

For whole exome sequencing of the same three samples, 0.1–0.5 µg of fragmented DNA extracted from paired tumor and normal FF tissues of three TRK-positive CRCs was prepared to construct libraries with the SureSelect Human All Exon Kit V5 (Agilent, Inc., USA) using the manufacturer’s protocol. Briefly, the qualified genomic DNA sample was randomly fragmented using Covaris followed by adapter ligation, purification, hybridization, and PCR. Captured libraries were subjected to an Agilent 2100 Bioanalyzer to estimate the quality and were loaded onto the NovaSeq 6000 platform (Illumina, CA, USA) according to the manufacturer’s recommendations.

### NGS data analysis for *NTRK* fusion detection

To identify *NTRK* fusions from the targeted RNA sequencing data of the 11 samples, sequencing reads were mapped to hg19 with STAR 2.7.3a,^29^ and processed with STAR-Fusion v1.9.0.^30^ STAR-Fusion uses the STAR-aligned reads to map junction and spanning reads to a junction annotation set. It then utilizes a chimeric junction file from STAR and produces a prediction file for gene fusion. The prediction file provides fused gene names, junction/spanning read counts, and breakpoint information. Analysis of the targeted RNA sequencing data was conducted in the central laboratory of the LOGONE Bio-Convergence Research Foundation.

From the whole transcriptome data of the three samples, *NTRK* fusion transcripts were identified using JAFFA, Arriba (https://github.com/suhrig/arriba), and FusionCatcher.^31^ The scheme of fusion transcripts was visualized using Arriba and potential functional consequences of fused protein were accessed using AGFusion and FusionHub,^32^ a database integrating 28 fusion-related datasets was used as an annotation database for known somatic fusions and *NTRK* fusions.

Using whole exome sequencing data, a novel *SFPQ*-*NTRK1* fusion, whose breakpoint was identified in whole transcriptome sequencing results, was validated by the Genome Rearrangement IDentification Software Suite (GRIDSS),^33^ a sensitive and specific genomic rearrangement detector using positional de Bruijn graph assembly and GeneFuse.^34^

### Fluorescence in situ hybridization

Fluorescence in situ hybridization (FISH) to detect DNA-level rearrangements of *NTRK1, NTRK2*, and *NTRK3* was performed in six TRK-positive SSL(D)s that were determined to be inappropriate for next-generation sequencing (NGS) analysis because of their limited, small amounts of tissues. FISH was conducted on the serrated epithelial area of each SSL or SSLD case using dual color break-apart probes for *NTRK1* (Empire Genomics, Williamsville, NY, USA), *NTRK2* (Empire Genomics), and *NTRK3* (Empire Genomics) according to the manufacturer’s protocols. FISH results were interpreted as follows: if 15% or more of counted nuclei in a case were positive for a break-apart signal or a red signal only, the case was considered positive for gene translocation of *NTRK1, NTRK2*, or *NTRK3*.^14^

### Statistical analysis

All statistical analyses and related data visualization were performed using GraphPad Prism version 9.0.0 (GraphPad Software, San Diego, CA, USA) or R version 4.0.2 (The R Foundation for Statistical Computing). Categorical variables were compared using the chi-square test or Fisher’s exact test. Continuous variables were compared using the Student’s t-test or Mann-Whitney U test. Statistical significance was determined at a two-sided *P*-value of < 0.05.

## RESULTS

### Landscape of pan-TRK expression in CRCs and premalignant colorectal lesions

We investigated pan-TRK IHC expression status in tissue samples encompassing a large series of CRCs (n = 441) and precursor lesions (n = 565). Representative pan-TRK IHC images are presented in Figure 1A, and the results of TRK positivity frequencies are summarized in Figure 1B. Notably, there were no TRK-positive cases among the MSS CRCs (n = 308), TSAs (n = 45), HPs (n = 100), CALGs (n = 150), and CAHGs (n = 115) (Figure 1). TRK-positive CRCs were 11 cases and were found exclusively in the MSI-high subset with *MLH1* promoter methylation (Figure 1). Among the various histological subtypes of premalignant colorectal lesions, only SSLDs (4 out of 23; 17%) and SSLs (5 out of 132; 4%) were determined to be TRK-positive cases (Figure 1). These findings strongly suggest that TRK protein overexpression, which is known to be mainly due to *NTRK* oncogenic rearrangements,^23, 24^ in benign or malignant colorectal epithelial tumors is closely associated with the serrated neoplasia pathway, a distinct multistep carcinogenesis pathway from SSLs to sporadic MSI-high and/or CpG island methylator phenotype-high (CIMP-high) CRCs.^17-19^

**Figure 1.**
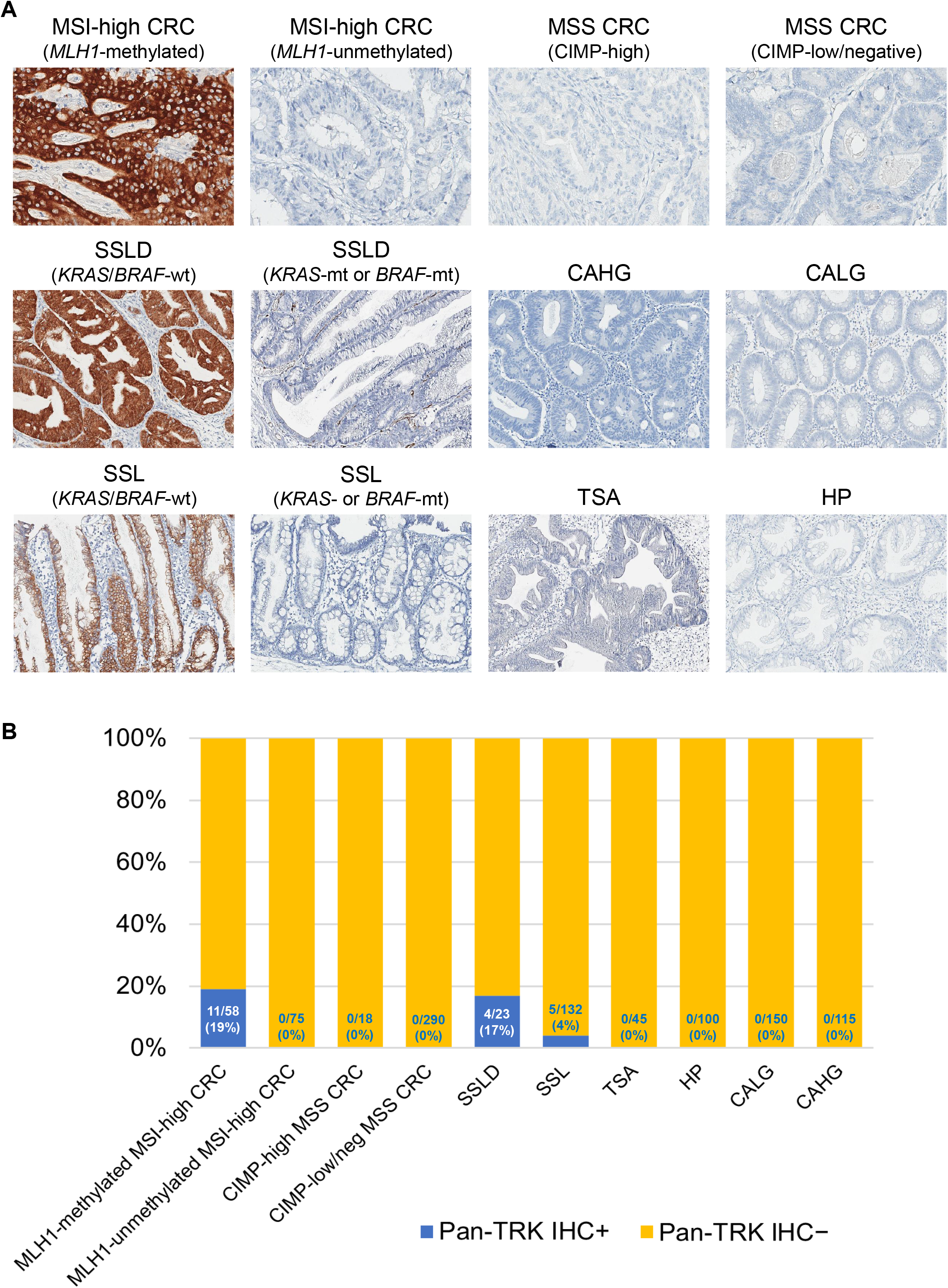
Pan-TRK expression status in CRCs and premalignant colorectal lesions. (A) Representative photomicrographs of pan-TRK IHC across various subtypes of CRCs and premalignant colorectal lesions. (B) Frequencies of pan-TRK IHC positivity in CRCs and premalignant colorectal lesions according to molecular or pathological subtypes Abbreviations: IHC, immunohistochemistry; CRC, colorectal cancer; MSI, microsatellite instability; MSS, microsatellite stable; CIMP, CpG island methylator phenotype; SSLD, sessile serrated lesion with dysplasia; wt, wild-type; mt, mutant-type; CAHG, conventional adenoma with high-grade dysplasia; CALG, conventional adenoma with low-grade dysplasia; SSL, sessile serrated lesion; TSA, traditional serrated adenoma; HP, hyperplastic polyp

### Characterization of *NTRK* fusion-positive CRCs

To confirm presence or absence of *NTRK* oncogenic fusions in the 11 CRCs showing pan-TRK IHC positivity, eight of the 11 TRK-positive MSI-high CRCs, which were available for using only FFPE tissues, were subjected to targeted RNA sequencing analysis, and three of the 11 TRK-positive MSI-high CRCs, which were available for using matched tumor and normal FF tissues, were analyzed by whole transcriptome and exome sequencing. *NTRK* fusions were detected in all 11 TRK-positive CRCs. The major clinicopathological and molecular characteristics of the 11 *NTRK* fusion-positive CRCs are summarized in Table 1. All 11 *NTRK* fusion-positive CRCs were commonly characterized by relatively old age (range 66–83), right-sided tumor location (ascending colon, hepatic flexure, or transverse colon; 100%), MSI-high (100%), CIMP-high (100%), *MLH1* promoter hypermethylation with loss of MLH1 IHC expression (100%), and absence of *KRAS*/*BRAF* mutations (100%) (Table 1). A total of six different partners of *NTRK1* or *NTRK3* fusions, including *TPM3*-*NTRK1* in three, *TPR*-*NTRK1* in two, *LMNA*-*NTRK1* in one, *SFPQ*-*NTRK1* in one, *EML4*-*NTRK3* in one, and *ETV6*-*NTRK3* in three, were identified in the 11 TRK-positive CRCs (Figures 2A-B and Table 1). Except for *SFPQ*-*NTRK1* fusion, the other five types of *NTRK* fusions were previously reported as oncogenic fusions in various tumor types, including CRCs (Figure 2A).^11-16^ However, the *SFPQ* gene has not been previously reported as a partner of *NTRK* fusions in human tumors, and thus, the *SFPQ*-*NTRK1* is a novel *NTRK* fusion that was first discovered in our present study. We confirmed that the *SFPQ-NTRK1* fusion was present as both RNA- and DNA-level structural alterations in the MSI-high CRC case (Figure 2B and Supplementary Figure S2). *SFPQ*-*NTRK1* fusion-positive CRC (Case No. MCRC-27) displayed moderate (2+) or strong (3+) intensity of pan-TRK IHC expression and was histologically characterized by sheets of poorly differentiated tumor cells with dense intratumoral lymphocytic infiltration, indicating a medullary carcinoma variant (Figure 2B and Table 1). The case also molecularly harbored MSI-high, CIMP-high, *MLH1* methylation, and *KRAS*/*BRAF* wild-type as other *NTRK* fusion-positive CRCs (Table 1).

**Table 1.**
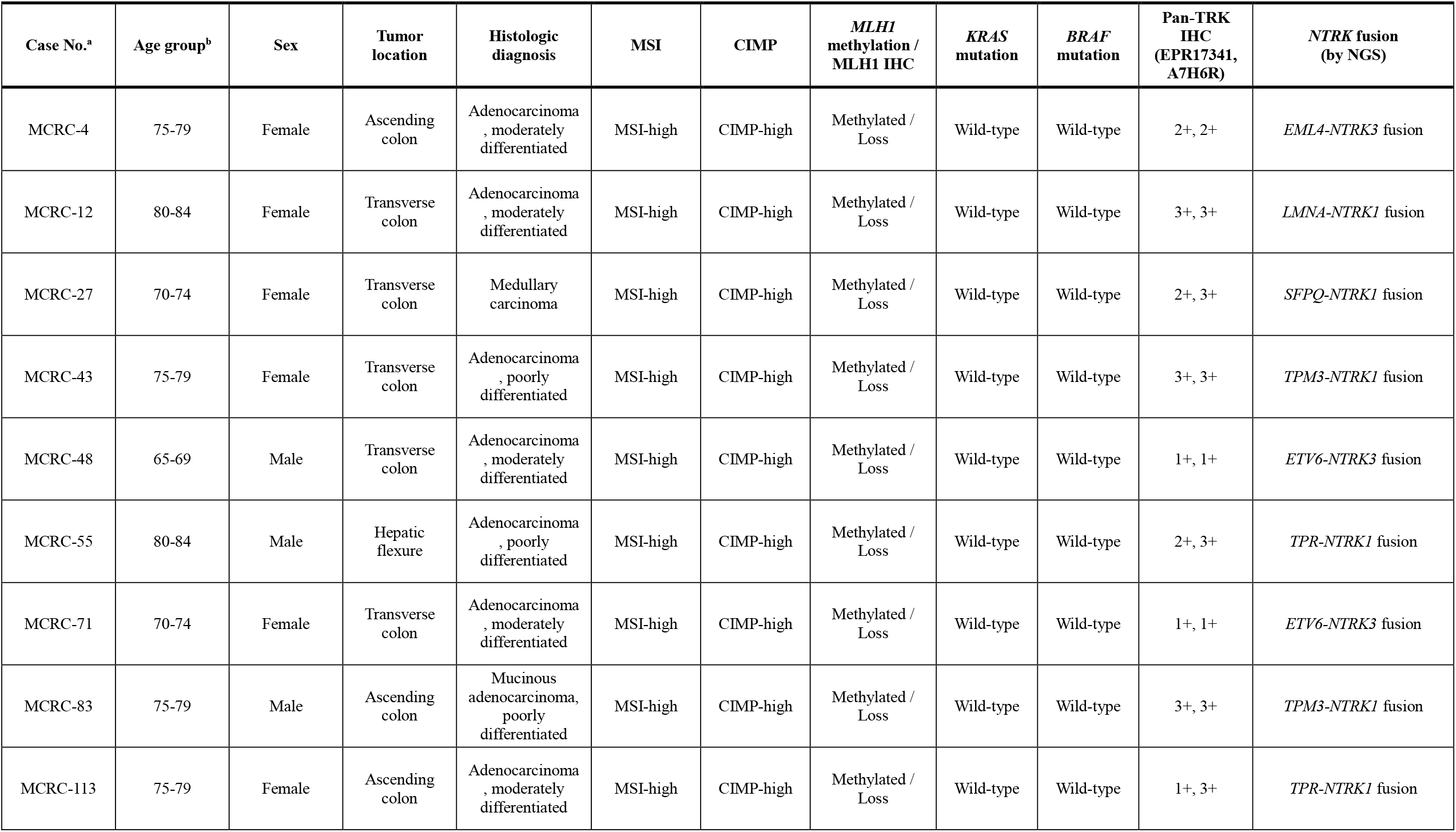
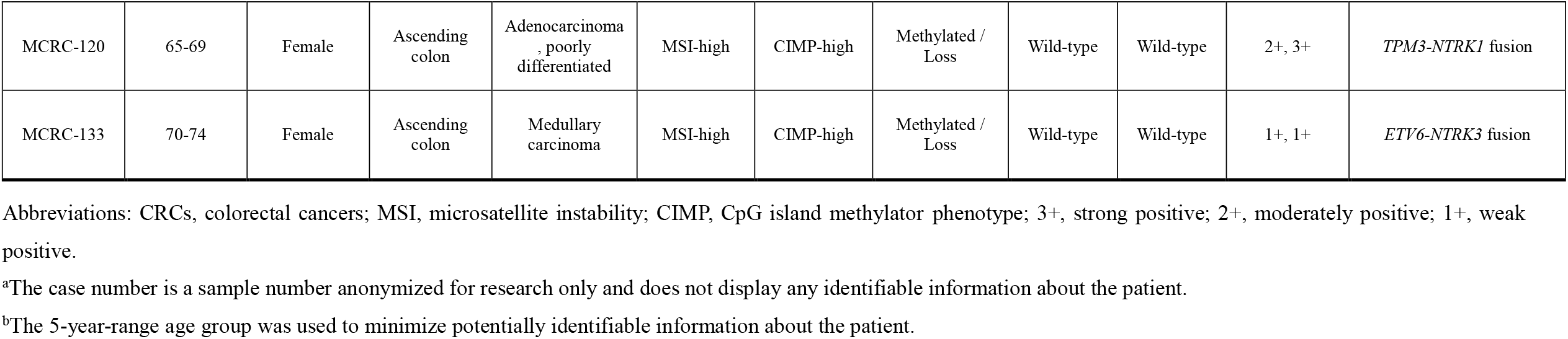
Summary of clinicopathological and molecular features of *NTRK* fusion-positive CRCs.

**Figure 2.**
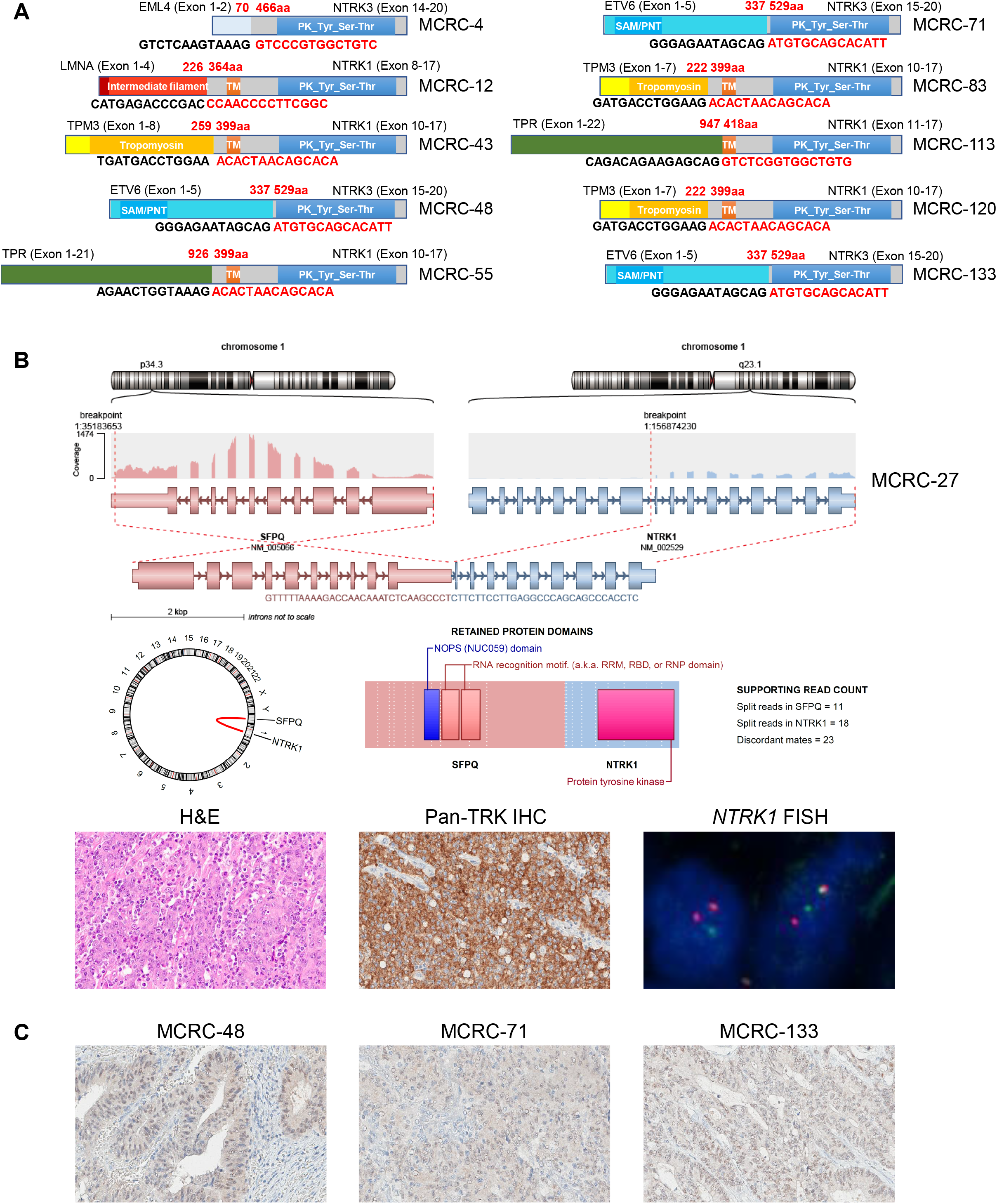
*NTRK* fusions and pan-TRK expression in TRK-positive MSI-high CRCs. (A) Schematic illustrations of *NTRK* fusions detected by targeted RNA sequencing or whole transcriptome sequencing in 10 TRK-positive MSI-high CRCs. All the fusions show appropriate direction and intact tyrosine kinase domain within the *NTRK* gene portion of their fusion transcripts. (B) A novel *SFPQ*-*NTRK1* fusion is identified by whole transcriptome sequencing in a TRK-positive MSI-high CRC (upper and middle). This tumor is histologically diagnosed as medullary carcinoma, which is characterized by sheets of poorly differentiated tumor cells with abundant tumor-infiltrating lymphocytes (lower left), shows moderate to strong intensity of pan-TRK IHC expression (lower middle), and is confirmed to have *NTRK1* translocation (break-apart signals by FISH) (lower right). (C) Photomicrographs of pan-TRK IHC in three MSI-high CRCs harboring an *ETV6*-*NTRK3* fusion. Note that weak nuclear and faint cytoplasmic expression patterns of pan-TRK are commonly observed in the three cases.

Interestingly, expression patterns of pan-TRK IHC in *NTRK* fusion-positive CRCs seemed to be different depending on the *NTRK* gene type or fusion partner gene. Eight of the 11 TRK-positive CRCs with a *TPM3*-*NTRK1, TPR*-*NTRK1, LMNA*-*NTRK1, SFPQ*-*NTRK1*, or *EML4*-*NTRK3* fusion showed easily identifiable moderate (2+) or strong (3+) staining intensity (Supplementary Figure S3 and Table 1); weak or faint expression (1+) of pan-TRK IHC was commonly observed in the three CRCs harboring an *ETV6*-*NTRK3* fusion (MCRC-48, MCRC-71, and MCRC-133) (Figure 2C and Table 1).

### Characterization of *NTRK* rearrangement-positive serrated colorectal lesions

We next investigated whether there were truly *NTRK* gene rearrangements in TRK-positive SSLDs and SSLs. NGS or FISH results for *NTRK* rearrangements in the nine TRK-positive SSL(D)s (four SSLDs and five SSLs) are summarized in Table 2 and Figure 3. The four TRK-positive SSLDs were subjected to targeted RNA sequencing analysis using RNA samples extracted from FFPE tissues, and *NTRK* oncogenic fusions were detected in two of the four SSLDs (*TPM3*-*NTRK1* and *ETV6*-*NTRK3*) (Figure 3A and Table 2). Identification of *NTRK* fusion by RNA sequencing failed in two SSLDs (Case No. SSLD-13 and SSLD-15) because of their poor RNA quality. Thus, these cases were re-analyzed by FISH using break-apart probes for the detection of *NTRK1, NTRK2*, and *NTRK3* rearrangements in tumor cell nuclei, and *NTRK3* and *NTRK1* translocations were successfully detected in the NGS-failed SSLDs (SSLD-13 and SSLD-15, respectively) (Figure 3B and Table 2). All four TRK-positive SSLDs commonly harbored CIMP-high and *KRAS*/*BRAF* wild-type (Table 2). However, in contrast to the *NTRK* fusion-positive CRCs found in this study, two TRK-positive SSLDs (SSLD-10 and SSLD-13) showed both MSS phenotype and intact MLH expression, although low-level *MLH1* promoter methylation was detected in these cases (Table 2 and Supplementary Figure S4).

**Table 2.** Summary of clinicopathological and molecular features of *NTRK* rearrangement-positive serrated lesions.

| Case No.<sup>a</sup> | Age group<sup>b</sup> | Sex | Tumor location | Histologic diagnosis | MSI | CIMP | *MLH1* methylation / *MLH1* IHC | *KRAS* mutation | *BRAF* mutation | Pan-TRK IHC (EPR17341, A7H6R) | *NTRK* rearrangement (by NGS or FISH) |
| --- | --- | --- | --- | --- | --- | --- | --- | --- | --- | --- | --- |
| SSLD-4 | 55-59 | Male | Ascending colon | Sessile serrated lesion with dysplasia (architecturally high-grade dysplasia) | MSI-high | CIMP-high | Methylated / Loss | Wild-type | Wild-type | 1+, 1+ | *ETV6-NTRK3* fusion |
| SSLD-10 | 65-69 | Female | Ascending colon | Sessile serrated lesion with dysplasia (intestinal type low-grade dysplasia) | MSS | CIMP-high | Methylated / No loss | Wild-type | Wild-type | 2+, 3+ | *TPM3-NTRK1* fusion |
| SSLD-13 | 60-64 | Female | Ascending colon | Sessile serrated lesion with dysplasia (serrated dysplasia) | MSS | CIMP-high | Methylated / No loss | Wild-type | Wild-type | 2+, 3+ | *NTRK3* translocation |
| SSLD-15 | 70-74 | Male | Descending colon | Sessile serrated lesion with dysplasia (architecturally high-grade dysplasia) and associated pT1 adenocarcinoma | MSI-high | CIMP-high | Methylated / Loss | Wild-type | Wild-type | 3+, 3+ | *NTRK1* translocation |
| SSL-10 | 50-54 | Female | Ascending colon | Sessile serrated lesion | MSS | CIMP-high | Unmethylated / No loss | Wild-type | Wild-type | 1+, 2+ (crypt base only) | *TPR-NTRK1* fusion |
| SSL-27 | 50-54 | Male | Ascending colon | Sessile serrated lesion | MSS | CIMP-high | Unmethylated / No loss | Wild-type | Wild-type | 1+, 1+ | *NTRK3* translocation |
| SSL-32 | 65-69 | Male | Hepatic flexure | Sessile serrated lesion | MSS | CIMP-high | Unmethylated / No loss | Wild-type | Wild-type | 1+, 1+ (crypt base only) | *NTRK3* translocation |
| SSL-52 | 60-64 | Female | Cecum | Sessile serrated lesion | MSS | CIMP-low | Unmethylated / No loss | Wild-type | Wild-type | 2+, 2+ | *NTRK1* translocation |
| SSL-55 | 70-74 | Male | Transverse colon | Sessile serrated lesion | MSS | CIMP-high | Methylated / No loss | Wild-type | Wild-type | 2+, 3+ | Undetermined |

Abbreviations: MSI, microsatellite instability; MSS, microsatellite stable; CIMP, CpG island methylator phenotype; 3+, strong positive; 2+, moderately positive; 1+, weak positive.
<sup>a</sup>The case number is a sample number anonymized for research only and does not display any identifiable information about the patient.
<sup>b</sup>The 5-year-range age group was used to minimize potentially identifiable information about the patient.

**Figure 3.**
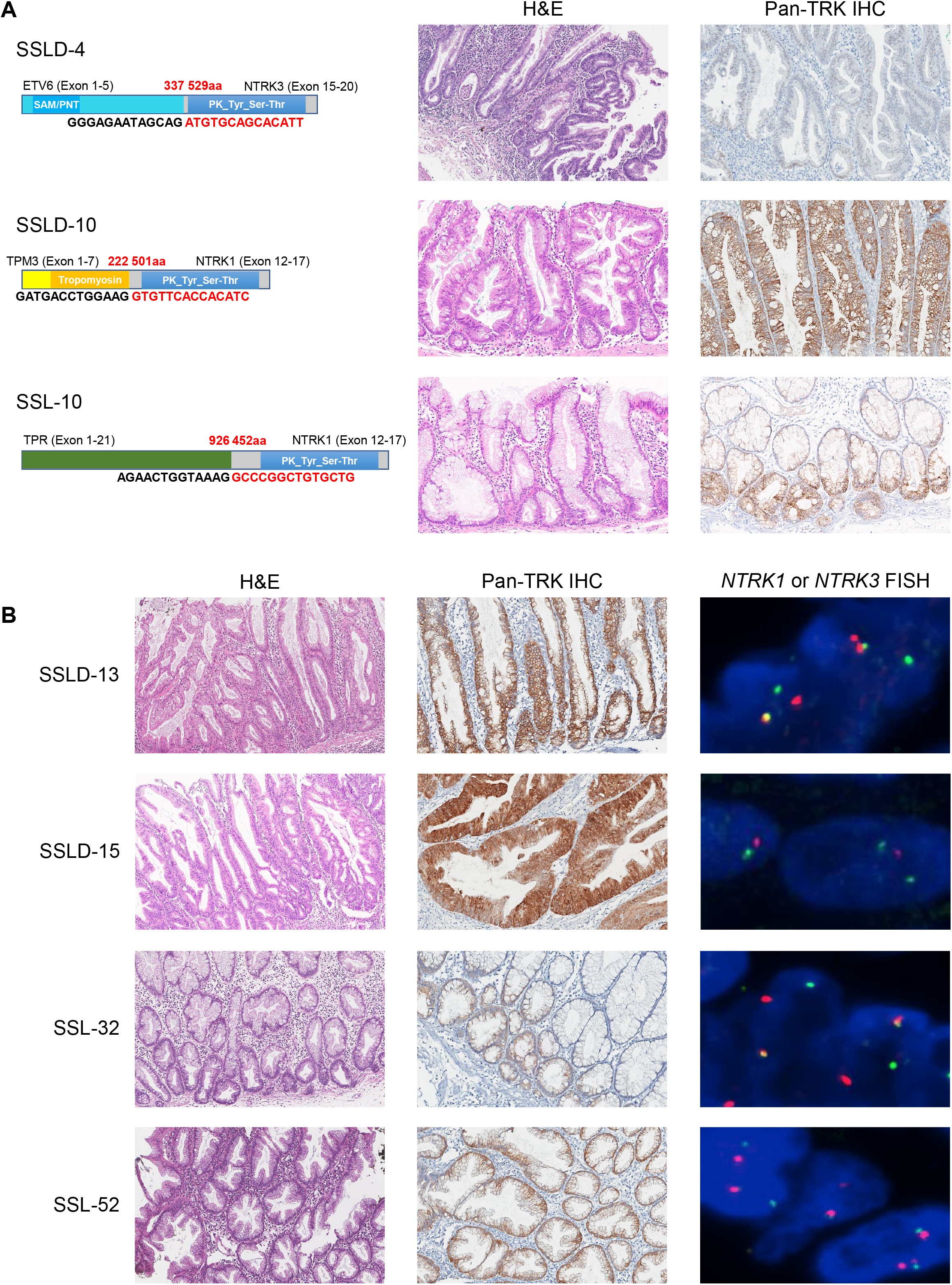
*NTRK* rearrangements and pan-TRK expression in TRK-positive SSL(D)s. (A) *NTRK* fusions detected by targeted RNA sequencing in three SSL(D)s. The SSLD-4 case is determined to harbor an *ETV6*-*NTRK3* fusion (upper left) and is histologically diagnosed as SSL with high-grade dysplasia/intramucosal carcinoma (upper middle). Note the weak intensity of pan-TRK IHC in the case (upper right). The SSLD-10 case is determined to harbor a *TPM3*-*NTRK1* fusion (middle left) and is histologically diagnosed as SSL with intestinal-type low-grade dysplasia (middle middle) and displays moderate to strong expression of pan-TRK IHC (middle right). The SSL-10 case is determined to harbor a *TPR*-*NTRK1* fusion (lower left) and is histologically diagnosed as SSL (lower middle). Note the characteristic expression pattern of pan-TRK IHC localized in crypt bases (lower right). (B) Four SSL(D)s are determined to have *NTRK1* or *NTRK3* translocation by FISH analysis. The SSLD-13 case is histologically diagnosed as SSL with serrated dysplasia (upper left) and shows moderate expression of pan-TRK IHC (upper middle) and displays *NTRK3* translocation (break-apart signals by FISH) (upper right). The SSLD-15 case is histologically diagnosed as SSL with architecturally high-grade dysplasia (middle-upper left) and shows strong expression of pan-TRK IHC (middle-upper middle) and displays *NTRK1* translocation (break-apart signals by FISH) (middle-upper right). The SSL-32 case is histologically diagnosed as SSL (middle-lower left) and shows weak expression of pan-TRK IHC (middle-lower middle) and displays *NTRK3* translocation (break-apart signals by FISH) (middle-lower right). Note the characteristic expression pattern of pan-TRK IHC localized in crypt bases (middle-lower middle). The SSL-52 case is histologically diagnosed as SSL (lower left), shows moderate expression of pan-TRK IHC (lower middle), and displays *NTRK1* translocation (break-apart signals by FISH) (lower right).

We also examined the presence or absence of *NTRK* gene rearrangements in the five TRK-positive SSLs without dysplasia. Because of the limitations of the remaining FFPE tissues after preceding extensive IHC/molecular studies, we failed to obtain the optimal quantity or quality of their RNA samples for NGS analysis in most SSLs. Therefore, targeted RNA sequencing was applicable only in one TRK-positive SSL, and instead, FISH analysis using break-apart probes for the detection of *NTRK1, NTRK2*, or *NTRK3* translocation was applied for the remaining four TRK-positive SSLs. *TPR*-*NTRK1* fusion was identified in the one sequenced SSL (SSL-10; Figure 3A and Table 2). FISH analysis revealed *NTRK1* translocation in one SSL (SSL-52), and *NTRK3* translocation was detected in two SSLs (SSL-27 and SSL-32) (Figure 3B and Table 2). However, in the remaining SSL (SSL-55), all *NTRK* FISH analyses failed to demonstrate identifiable fluorescence signals, probably due to poor tissue quality (Table 2). Although all five TRK-positive SSLs commonly demonstrated the absence of *KRAS*/*BRAF* mutations, one case (SSL-52) harbored CIMP-low instead of CIMP-high, and four cases (SSL-10, SSL-27, SSL-32, and SSL-52) did not show *MLH1* promoter methylation or loss of MLH1 expression (Table 2 and Supplementary Figure S5).

### Intralesional heterogeneity of pan-TRK expression in SSLs and its morphological correlations

We further noted the intralesional heterogeneity of pan-TRK IHC expression in SSLs and discovered two important patterns from these observations (Figures 4A-B). First, in two SSLs (SSL-10 with *TPR*-*NTRK1* fusion and SSL-32 with *NTRK3* translocation), pan-TRK IHC expression was localized mainly at the basal crypt epithelium of the SSLs (Figures 3A-B and 4A). We carefully re-evaluated *NTRK* FISH encompassing full longitudinal portions of crypts of the two SSLs and found that *NTRK* rearrangement was commonly observed in both the upper and lower portions of the epithelium of the SSLs, indicating clonal involvement of DNA-level *NTRK* rearrangement throughout serrated crypts despite their heterogeneous TRK expression (Figure 4A). Furthermore, although these two cases were originally classified as SSLs without dysplasia, mild nuclear atypia such as relatively enlarged nuclei with occasional distinct nucleoli, suggesting early morphological changes into serrated dysplasia, were observed only in the basal crypt epithelial cells of an SSL (SSL-10) (Figure 4A). Based on this finding, we propose a hypothetical model explaining the sequential mechanism of *NTRK* fusion-related dysplastic changes that begin at crypt bases in SSLs (Figure 4C).

**Figure 4.**
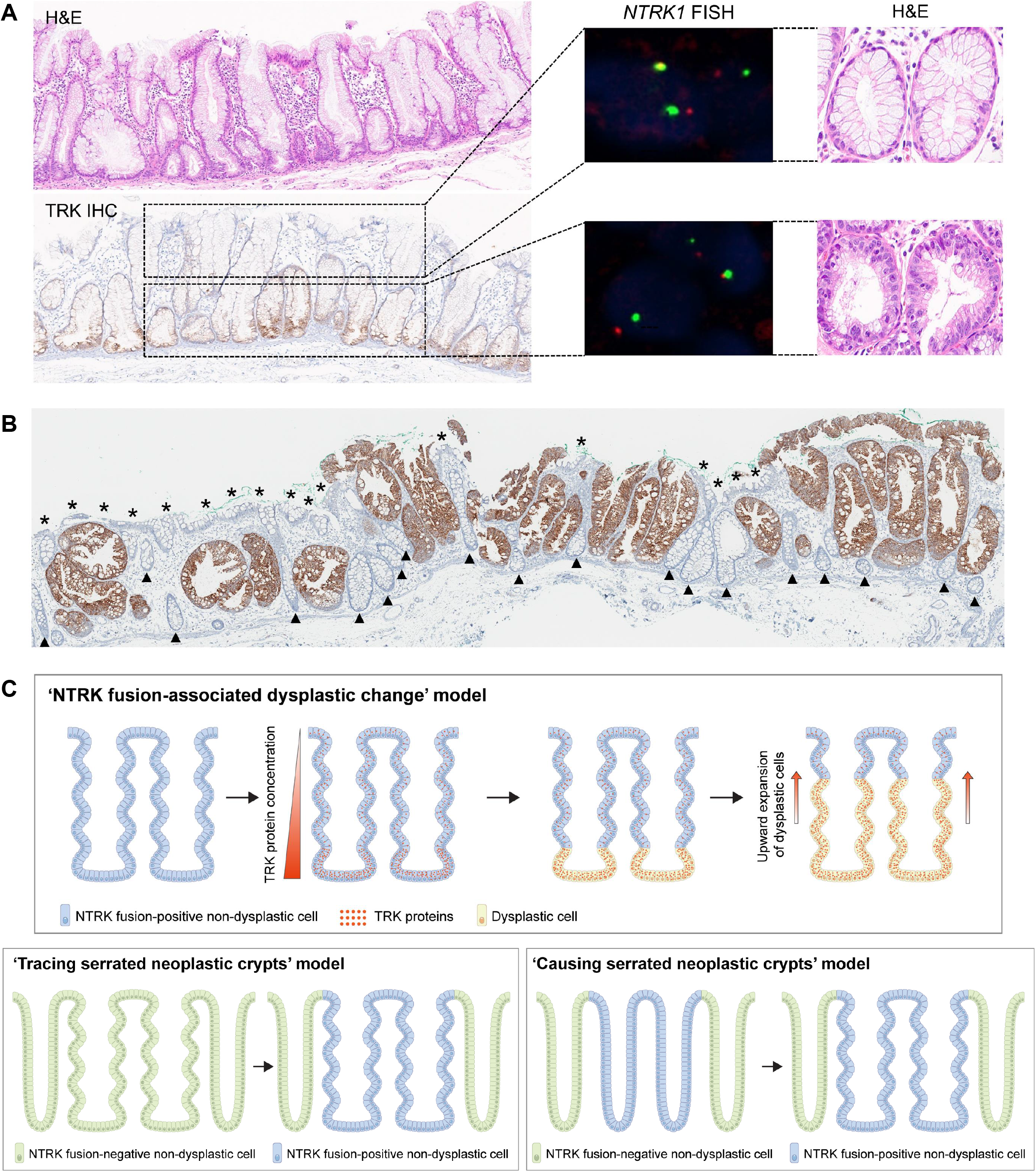
TRK expression heterogeneity in SSLs and its correlation with morphological features. (A) Pan-TRK IHC expression is localized in the base to lower third of crypt epithelium within an SSL harboring a *TPR*-*NTRK1* fusion (Case No. SSL-10). By contrast, FISH analysis using *NTRK1* break-apart probes demonstrates presence of *NTRK1* translocation in epithelial cells of both upper and lower areas of crypts in the SSL. In high-power views of an H&E-stained section, relatively enlarged, vesicular nuclei with visible nucleoli, suggesting early morphological changes toward serrated dysplasia, are observed only at the basal crypt epithelium, but not at the upper crypt epithelium, in the SSL. (B) Pan-TRK IHC staining is positive selectively in architecturally distorted serrated crypts within an SSLD (Case No. SSLD-10). Note the absence of TRK expression in both non- or less-serrated crypts (arrowheads) and the non-dysplastic surface epithelium (asterisks) within the SSLD. (C) Conceptual models of *NTRK* fusion-related morphological changes within an SSL. (Upper) A ‘bottom-up’ model for *NTRK* fusion-related dysplastic change consists of the following steps: (1) clonal expansion of *NTRK* fusion-positive cells along the epithelium of distorted serrated crypts, (2) localization of *NTRK* fusion-induced TRK protein overexpression at basal crypt cells due to an uncertain mechanism, (3) initiation of cytological dysplasia at basal crypt cells, and (4) upward spread of both TRK overexpression and morphological dysplasia along distorted serrated crypts. (Lower) Two hypothetical models elucidating the relationship between *NTRK* fusions and distorted serrated crypts within an SSL: (1) preceding formation of distorted serrated crypts and subsequent spread of *NTRK* fusion-positive cells selectively along the distorted serrated crypts (‘tracing serrated neoplastic crypts’ model); and (2) preceding expansion of *NTRK* fusion-positive epithelial cells along non-distorted serrated crypts and subsequent morphogenesis of distorted serrated crypts selectively along the *NTRK* fusion-positive crypts (‘causing serrated neoplastic crypts’ model).

As a second finding, we revealed that TRK could be discontinuously expressed in the surface epithelium and stained selectively in architecturally distorted serrated crypts, a histological hallmark defining SSLs as premalignant neoplastic lesions and distinguishing SSLs from HPs (Figure 4B). This spatial concordance between TRK expression and neoplastic serrated crypts in SSLs can raise two possibilities regarding the mechanistic relationship between *NTRK* rearrangement and SSL-specific morphogenesis as follows: (1) clonal spread of *NTRK* fusion-positive cells may selectively involve pre-existing distorted serrated crypts (‘tracing serrated neoplastic crypts’) within an SSL; or (2) clonal spread of *NTRK* fusion-positive cells may selectively promote the morphological change of non-serrated crypts into distorted serrated crypts (‘causing serrated neoplastic crypts’) within and around an SSL (Figure 4C). Both hypotheses were supported by detailed histological features, including a fully serrated crypt showing focal weak staining of pan-TRK IHC (Supplementary Figure S6) or a mixed serrated and non-serrated crypt showing staining of pan-TRK IHC only at the intra-crypt serrated morphological area (Supplementary Figure S7).

### Comparison of age distributions of serrated pathway-associated molecular alterations

Finally, we investigated the age distribution patterns of SSLs, SSLDs, and sporadic MSI-high CRCs in the context of serrated pathway-associated molecular alterations, including *BRAF* mutation, *KRAS* mutation, *NTRK* fusion, CIMP-high, and *MLH1* methylation (Figure 5). As expected, average ages sequentially increased from SSLs to sporadic MSI-high CRCs, with statistical significance (Figure 5A). We also compared age differences between molecular alteration-positive SSL(D)s and sporadic MSI-high CRCs and found that there were significant differences in average ages between each molecular alteration-positive SSL(D)s and sporadic MSI-high CRCs (Figure 5A). In contrast to other molecular alterations that were detected only in SSLs of patients aged ≥ 50 years, *BRAF* mutation was found in younger age-onset SSLs. Notably, both the average age interval and the earliest age interval between serrated precursors and MSI-high CRCs were the longest in *BRAF* mutation-positive tumors (19 years and 21 years, respectively), whereas they were shortest in *MLH1*-methylated tumors (9 years and less than 1 year, respectively) (Figure 5A).

**Figure 5.**
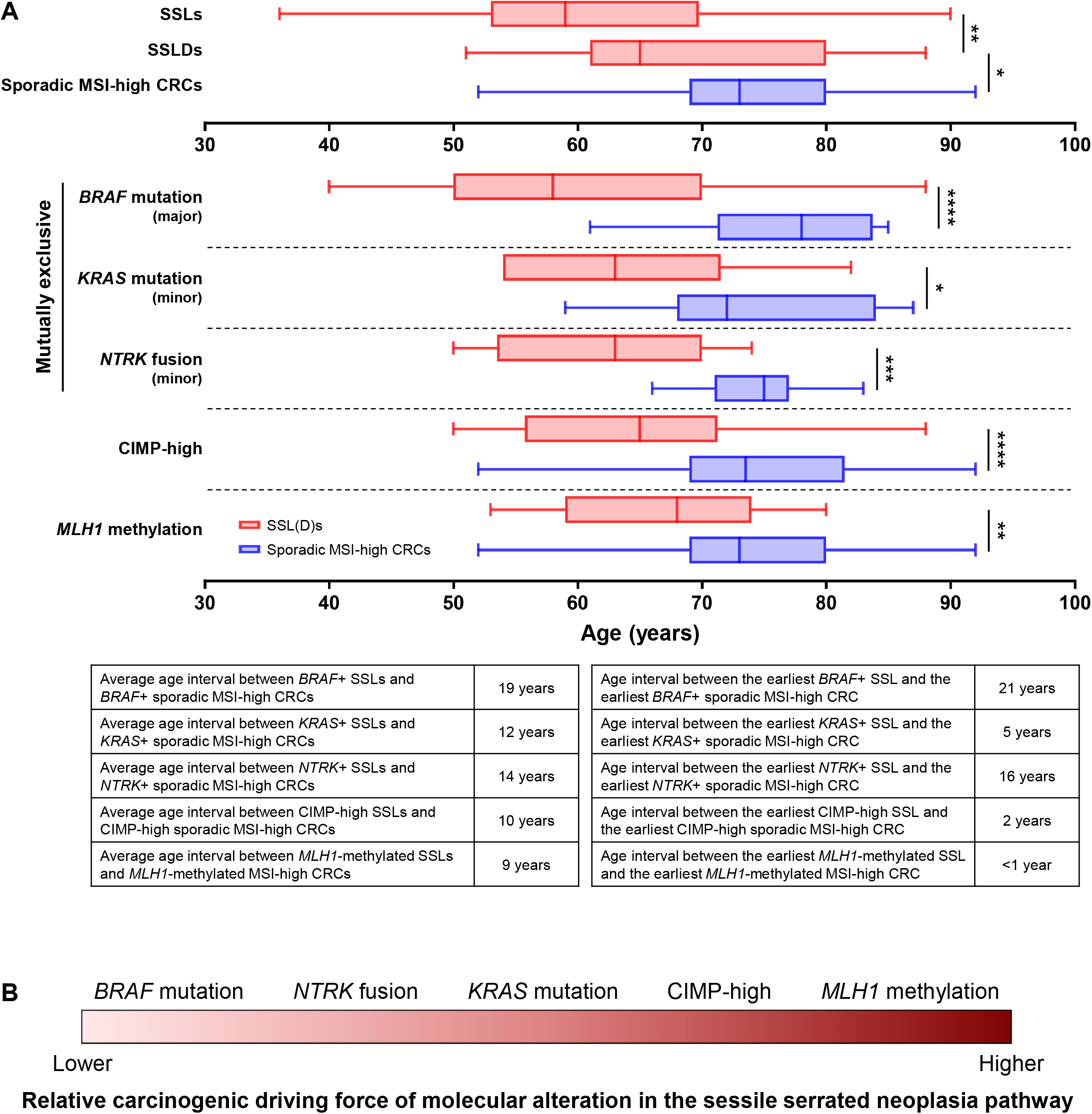
Age distribution patterns of molecular alterations in the sessile serrated neoplasia pathway. (A) Box-whisker plots showing age distributions of sporadic MSI-high CRCs and their precursors, including SSLs and SSLDs, with or without consideration of molecular alterations. Upper column plots demonstrate age distributions of total SSLs (n = 132), SSLDs (n = 23), and sporadic MSI-high CRCs (n = 58). Middle column plots display age distributions of each molecular alteration-positive tumors to compare time intervals between each molecular alteration-positive SSL(D)s and sporadic MSI-high CRCs. Lower column tables summarize age intervals between each molecular alteration-positive SSL(D)s and sporadic MSI-high CRCs. A central line in a box, a box length, and bilateral whiskers indicate a median, an interquartile range, and a minimum to maximum range, respectively. (****, *P* < 0.0001; ***, *P* < 0.001; **, 0.001 ≥ *P* < 0.01; *, 0.01 ≥ *P* < 0.05) (B) A schematic model comparing relative oncogenic driving force of each molecular alteration in the sessile serrated neoplasia pathway. This estimation is inferred from the above age interval data of each molecular alteration and does not consider potential combined effects of the molecular alterations.

Collectively, based on these findings, we could infer the relative carcinogenic driving force of each molecular alteration in the sessile serrated neoplasia pathway (Figure 5B). Because the time intervals between *BRAF*-mutated SSLs and *BRAF*-mutated MSI-high CRCs were the longest, the immediate risk of malignant transformation in young age-onset *BRAF*-mutated SSLs may be lower than that in other molecular alteration-positive SSLs. However, since relatively short age intervals were observed between SSLs and MSI-high CRCs with CIMP-high or *MLH1* methylation, these epigenetic alterations may be associated with relatively rapid malignant changes in SSLs. Based on a similar estimation, the carcinogenic driving power of *NTRK* fusions may be intermediate between that of *BRAF* mutation and that of CpG island methylation in the sessile serrated neoplasia pathway (Figure 5B).

## DISCUSSION

In our primary CRC tissue cohorts consisting of 133 MSI-high CRCs and 308 MSS CRCs, TRK-positive cases were found only in MSI-high CRCs but not in MSS CRCs (Figure 1). All 11 TRK-positive CRCs were confirmed to harbor *NTRK* fusions by targeted RNA or whole transcriptome sequencing analyses. There were six types of *NTRK* fusions (*TPM3*-*NTRK1, TPR*-*NTRK1, LMNA*-*NTRK1, SFPQ*-*NTRK1, EML4*-*NTRK3*, and *ETV6*-*NTRK3*) in the 11 TRK-positive CRCs (Table 1 and Figure 2), and all of the fusions, except for *SFPQ*-*NTRK1*, had been previously discovered as oncogenic fusions in CRCs.^11-16^ Interestingly, multiple research groups recently reported unique associations between oncogenic fusions and the underlying molecular features of CRCs.^11-15^ The studies have commonly described that major oncogenic fusions, including *NTRK, ALK, RET*, and *BRAF* fusions, were detected only in a sporadic MSI-high subset with *MLH1*-methylation and *KRAS*/*BRAF* wild-type.^11-13^ These characteristics are also confirmed by our present study: in detail, all of the 11 *NTRK* fusion-positive CRCs had common molecular features, including MSI-high, CIMP-high, *MLH1* promoter methylation, MLH1 expression loss, and absence of *KRAS*/*BRAF* mutations (Table 1). Because activating mutation in *KRAS* or *BRAF* gene is regarded as one of the strongest oncogenic drivers in human cancers, including CRC,^43, 44^ the absence of both *KRAS* and *BRAF* mutations in a tumorigenesis pathway generally requires an alternative powerful oncogenic driver such as mutation, amplification, or fusion of other major oncogenes. Thus, it is quite reasonable that there have been consistent findings that *NTRK* oncogenic fusions occur specifically in a *KRAS*/*BRAF* wild-type subset among CRCs. A recent study also supported the fact that *NTRK* fusions are enriched in tumors without canonical driver mutations.^9^

Because *NTRK* fusion-positive CRCs have been found only in a sporadic MSI-high subset (*MLH1*-methylated subset) of CRCs, it can be inferred that clinicopathological features of *NTRK* fusion-positive CRCs may follow those of sporadic MSI-high CRCs. Indeed, according to our data, relatively old age (average 75; range 66–83), female predominance (73%), right-sided colonic tumor location (100%), and relatively frequent poorly differentiated histology including medullary variant (55%) were clinicopathological characteristics of *NTRK* fusion-positive CRCs (Table 1), all of which are also known to be closely associated with sporadic MSI-high CRCs.^35^ We additionally compared various clinicopathological and immunological features between *NTRK* fusion-positive (n = 11) and -negative (n = 47) subsets of the 58 sporadic MSI-high CRCs, but there were no significantly different factors except for a few molecular factors such as *KRAS*/*BRAF* mutations between these subgroups (Supplementary Table S1), suggesting that *NTRK* fusion itself may have little effect on differential clinicopathological phenotypes within sporadic MSI-high CRCs.

The most important achievement of our study was that it was the first to confirm that *NTRK* fusions occur exclusively in SSL(D)s among various precursors of CRCs (Figure 1). This result is quite plausible because SSLs are known to be major precursors of sporadic *MLH1*-methylated MSI-high CRCs, a CRC subtype that has been reported to specifically harbor *NTRK* fusions according to our present and others’ previous data.^11-13^ Consistent with the findings in CRCs (Table 1), there was also an absence of *KRAS*/*BRAF* mutations in all the *NTRK*-rearranged SSL(D)s (Table 2). However, interestingly, not all the *NTRK*-rearranged SSL(D)s harbored CIMP-high and/or *MLH1* methylation (Table 2), which are essential molecular factors for sporadic MSI-high CRCs. These features can induce two inferences: (1) *NTRK* fusions in SSLs may start before extensive CpG island methylation occurs in SSLs; (2) dysplastic changes in some *NTRK* fusion-positive SSLs may be independent of CIMP/MLH1 status. As our present study is an observational study using human tissue samples, there is a limitation to understanding why *NTRK* fusions occur exclusively in SSLs and not in conventional adenomas. Hence, additional *in vitro* and *in vivo* experimental studies should be conducted to unveil the underlying molecular mechanisms of the pathway specificity of oncogenic fusions in colorectal carcinogenesis.

Based on our finding that the proportion of *NTRK*-rearranged tumors progressively increased from benign to malignant lesions along the serrated neoplasia pathway (4% in SSLs, 17% in SSLDs, and 19% in sporadic MSI-high CRCs; Figure 1B), it can be inferred that *NTRK* fusions may undergo positive selection during the progression of serrated neoplasia and may be one of the critical molecular drivers of dysplastic and/or malignant transformation of SSLs. In other words, TRK-positive SSLs may be considered to be at an increased risk for advanced lesions. Thus, further validation studies are necessary to evaluate the potential utility of pan-TRK IHC positivity as an indicator of high-risk lesions among non-dysplastic SSLs. If pan-TRK IHC is determined to be helpful for selecting high-risk SSLs as other currently used risk factors such as morphological dysplasia, lesion size, and lesion multiplicity,^19, 36, 37^ it will allow us to perform more precise surveillance and management of SSLs. Moreover, other IHC items that have been known to be useful for detecting oncogenic fusions, such as ALK IHC for *ALK* fusions,^38^ may also be predictive factors for high-risk SSLs. In fact, the specific presence of *ALK* fusions in sporadic MSI-high CRCs has been reported in other studies,^11-13^ and we also found an ALK-positive case among the 132 SSLs (Supplementary Figure S8). Similar to *NTRK* fusions, the potential dependency of *ALK* fusions on the serrated neoplasia pathway in the colorectum should be further confirmed.

Another interesting finding in our data was the intralesional heterogeneity of pan-TRK expression and its correlation with early dysplastic change or distorted serrated crypts in SSLs (Figure 4). These features provide important insights into the initiation and spreading patterns of *NTRK* fusion-associated dysplastic and architectural changes within an SSL. First, the initiation point of molecular alterations over pathogenic threshold values within an SSL may be crypt base cells, according to our finding that TRK positivity was localized in the crypt base epithelium of some SSLs (Figure 4A). A previous theory explaining the mechanisms of the morphological change into architecturally distorted serrated crypts, a histological hallmark of SSLs, was that the location of an epithelial proliferation zone might be abnormally shifted from the crypt base to the lateral or surface side of the crypt.^39^ However, our findings indicate that molecular aberrations beyond a certain threshold for dysplastic change, represented by overexpression of TRK protein, may be prone to begin at basal crypt cells within an SSL, although DNA-level changes such as *NTRK* fusion have already been present throughout serrated crypts (Figures 4A and 4C). The potential ‘bottom-up’ feature of molecular change and its associated dysplastic change in SSLs is in contrast to the canonical ‘top-down’ feature of dysplastic change typically observed in intestinal-type dysplasia of conventional adenomas. The ‘bottom-up’ hypothesis of molecular-dysplastic change in SSLs can also be supported by our finding that loss of MLH1 expression was partially observed in the lower portion of crypts within an *MLH1*-methylated SSL (Supplementary Figure S9).^18, 19^ Moreover, recent efforts identified several non-conventional dysplasia patterns in the gastrointestinal tract, and among them, basal crypt dysplasia in Barrett esophagus, basal gland dysplasia in the stomach, and crypt cell dysplasia in inflammatory bowel disease are representatives of dysplasia that begins at the mucosal base areas.^40, 41^ Thus, TRK expression-associated early dysplastic features confined to crypt bases found in some of our SSL cases may be interpreted as another example of non-conventional dysplasia patterns. Another insight can be obtained from the finding that TRK expression was observed selectively in typical distorted serrated crypts, but not in non-serrated or less-serrated crypts, within SSLs (Figure 4B). This characteristic supports two hypothetical sequence models: a ‘tracing serrated neoplastic crypts’ model and a ‘causing serrated neoplastic crypts’ model (Figure 4C). Both models are plausible because a few serrated crypts with faint or negative TRK expression were occasionally observed at the periphery of TRK-positive SSLs (supporting the ‘tracing serrated neoplastic crypts’ model; Supplementary Figure S6), whereas a few mixed morphological crypts showed TRK positivity only at the serrated portion within the crypt (supporting the ‘causing serrated neoplastic crypts’ model; Supplementary Figure S7). To understand the precise relationship between molecular alterations and morphological changes during the pathogenesis of SSLs, further studies should be conducted.

One of the noticeable findings in our study was the identification of the *SFPQ* gene as a novel partner for *NTRK* oncogenic fusion. The *SFPQ* gene is known to be one of the major partners for *TFE3* fusions in Xp11 translocation renal cell carcinomas or perivascular epithelioid cell tumors,^45^ but to the best of our knowledge, it has not been reported to be a partner for *NTRK* fusions. The *SFPQ-NTRK1* fusion found in an MSI-high CRC from our study cohort can be acknowledged as a functionally oncogenic fusion based on the following evidence:

1. The *SFPQ-NTRK1* fusion transcripts contained the intact kinase domain of *NTRK1* with the tyrosine docking site downstream of the fusion (Figure 2B), which is a key characteristic of functionally oncogenic *NTRK* fusions.
2. The *SFPQ* gene, a partner gene upstream, has a coiled-coil domain that is required for TRK activation. The SFPQ protein encoded by the *SFPQ* gene is in the Drosophila behavior/human splicing (DBHS) protein family, which contains four domains: RNA recognition motifs 1 and 2, a nonA/paraspeckle domain, and a coiled-coil domain.^46^ The coiled-coil domain is a domain that allows protein-protein interaction and is frequently included in the upstream partner gene of *NTRK* oncogenic fusions.^47^ It is also required for the ligand-independent activation of various kinase fusions, including downstream kinase domains.^47, 48^ *NTRK* fusions are known to frequently have upstream partner genes with a coiled-coil domain, such as *MPRIP, TPM3, TPR, TFG, ARHGEF2, LMNA, SQSTM1, TRIM63*, and *PPL* for *NTRK1* fusions, *TRIM24, PAN3*, and *SQSTM1* for *NTRK2* fusions, and *TPM4, TFG, MYO5A, MYH9, STRN3, STRN*, and *EML4* for *NTRK3* fusions.^47, 49^
3. Using whole exome sequencing analysis, we confirmed that the *SFPQ-NTRK1* fusion was also present as a DNA-level structural alteration in the MSI-high CRC case with *SFPQ-NTRK1* fusion transcripts detected by RNA sequencing (Supplementary Figure S2).
4. The *SFPQ-NTRK1* fusion-positive case demonstrated moderate to strong positive expression of pan-TRK IHC in tumor cells (Figure 2B). As described in detail below, pan-TRK IHC positivity is a reliable indicator of the presence of *NTRK* oncogenic fusions in tumor tissues.

Several previous investigations have proven that pan-TRK IHC is suitable for screening of *NTRK* oncogenic fusions across various solid tumors.^21-24^ Solomon et al. reported that the sensitivity and specificity of pan-TRK IHC for detecting *NTRK* fusions in CRCs were 87.5% and 100%, respectively.^22^ Our present study also confirmed that the positivity of pan-TRK IHC in colorectal tumors was well matched with the presence of *NTRK* rearrangements identified by NGS or FISH, with a positive predictive value of 100%. However, we simultaneously found an important pitfall of pan-TRK IHC, its differential staining intensities depending on the partner genes of *NTRK* fusions, in colorectal tumors. According to our data, all four *ETV6*-*NTRK3* fusion-positive colorectal tumors, including three CRCs and one SSLD, commonly showed weak (1+) intensities of pan-TRK IHC expression, regardless of the tested antibody clones (EPR17341 or A7H6R) (Figures 2C and 3A). In particular, among the cases, pan-TRK staining intensities in two CRCs (MCRC-71 and MCRC-133) with an *ETV6*-*NTRK3* fusion were too faint to be easily recognized even though experienced pathologists participated in the evaluation (Figure 2C). This finding suggests that pan-TRK IHC expression may be misinterpreted as negative in colorectal tumors with an *ETV6*-*NTRK3* fusion. Indeed, it has been suggested that the staining characteristics of pan-TRK IHC in *NTRK* fusion-positive tumors vary depending on fusion partners or tumor/tissue types.^22, 50, 51^ According to recent reports, *ETV6*-*NTRK3* fusion-positive tumors generally demonstrate nuclear to cytoplasmic staining of pan-TRK IHC in their tumor cells, and occasionally some *ETV6*-*NTRK3* fusion salivary gland tumors show focal weak staining of pan-TRK IHC, similar to our cases.^21, 22, 50^ Although the overall sensitivity of pan-TRK IHC for the detection of *NTRK1* or *NTRK2* fusion tumors was reported to be high (96% and 100%, respectively), the sensitivity of pan-TRK IHC for *NTRK3* fusion tumors was relatively low (79%).^22^ Therefore, in colorectal tumors, the potential limited sensitivity of pan-TRK IHC for *NTRK3* fusion detection should be considered and further investigated.

Although it is known that the *BRAF* V600E mutation is a predominant molecular alteration in SSL(D)s,^26, 42^ our results regarding age intervals between SSLs and sporadic MSI-high CRCs with or without specific molecular alterations indicate that the relatively rare *NTRK* fusions appear to be a more powerful driver for malignant transformation of SSLs than *BRAF* mutation (Figure 5). Because our study is a retrospective observational study, our results have intrinsic limitations in confirming these inferred conclusions, and well-designed longitudinal follow-up studies are necessary. However, in addition to the age distribution data, other clinicopathological and molecular features also support our reasoning as follows: compared with TRK-negative SSLs, TRK-positive SSLs were significantly associated with larger lesion size (≥ 10 mm, 100%; *P* = 0.011) and CIMP-high status (80%; *P* = 0.014) (Supplementary Table S2), which are known pathological and molecular risk factors for progression of SSLs into advanced neoplasms. These findings collectively indicate that *NTRK* fusions may occur preferentially in higher-risk SSLs in relatively older (≥ 50 years) patients. Thus, the potential clinical application of pan-TRK IHC as a screening tool for detecting high-risk SSLs should be considered in further clinical studies.

There are several limitations to this study because it was a retrospective tissue-based observational study with limited sample size. First, in contrast to *NTRK1* or *NTRK3* fusions, no *NTRK2* fusions were detected by NGS or FISH analysis in our TRK-positive samples. In detail, among the 20 TRK-positive colorectal tumors, including 11 CRCs and 9 SSL(D)s, *NTRK1* rearrangements were confirmed in seven CRCs and four SSL(D)s, and *NTRK3* rearrangements were found in four CRCs and four SSL(D)s (Tables 1 and 2). RNA- or DNA-level *NTRK* rearrangements in the remaining SSL was not identified by either NGS or FISH analyses (Table 2). Accordingly, it cannot be excluded that the undetermined case may have an *NTRK2* fusion. In addition, it cannot be ruled out that pan-TRK IHC may not be suitable for the detection of *NTRK2* fusions in colorectal tumors. However, in fact, *NTRK2* fusions have been known to be relatively rare in overall tumors compared to *NTRK1* or *NTRK3* fusions,^1^ and *NTRK2* fusions have not been reported in CRCs, except for one case (*MUC2*-*NTRK2* fusion) discovered by a recent investigation.^14^ Moreover, according to large data, the sensitivity of pan-TRK IHC for the detection of *NTRK2* fusions in various solid tumors was reported to be 100%.^22^ Collectively, the absence of *NTRK2* fusions in our study cohort may not be based on analytical errors, but rather it may be based on the extreme rarity of *NTRK2* fusions in colorectal tumors.

Another limitation of our study was that there was no TRK positivity in MSS/CIMP-high CRCs. Considering that SSLs are regarded as major precursors of CIMP-high CRCs with or without *MLH1* methylation,^17-19^ theoretically, *NTRK* fusions are expected to be found in MSS/CIMP-high (*MLH1*-unmethylated) CRCs as well as MSI-high/CIMP-high (*MLH1*-methylated) CRCs. Among the 18 cases determined to be MSS/CIMP-high CRCs in our study cohort, there was only one tumor satisfying *KRAS*/*BRAF* wild-type, indicating that the number of candidates for the exploration of *NTRK* fusions in MSS/CIMP-high CRCs was too small. Interestingly, we revealed that *NTRK* rearrangements were present in SSL(D)s without *MLH1* methylation (Table 2). Among them, two *NTRK*-rearranged SSLDs (SSLD-10 and SSLD-13) were confirmed to harbor MSS and CIMP-high (Table 2). These cases indicate that dysplastic change can start in *NTRK* fusion-positive SSLs even under the MSS condition, suggesting the possibility of the presence of *NTRK* fusions in MSS/CIMP-high CRCs. In fact, until recently, a few MSS CRC cases were reported to have *NTRK* fusions.^11, 14, 15^ We strongly suggest that all the MSS/*NTRK*-positive CRCs found in previous studies might be SSL-derived MSS/CIMP-high tumors. Although it seems that *NTRK* fusions are relatively rare in MSS/CIMP-high CRCs compared with sporadic MSI-high CRCs, it should be considered that *NTRK* fusion can also be present in MSS CRCs.

In summary, to the best of our knowledge, our study is the first to present the full clinicopathological and molecular landscape of pan-TRK expression and *NTRK* rearrangements from benign to malignant colorectal tumors encompassing serrated and conventional neoplasia pathways. *NTRK* oncogenic fusions are found specifically in sessile serrated pathway-associated tumors without *KRAS*/*BRAF* mutations, including SSLs, SSLDs, and sporadic MSI-high CRCs. *NTRK* fusions are a relatively early event that can occur morphologically prior to full dysplastic change and molecularly prior to MSI-H development in SSLs.

## Supporting information

Supplementary Table S3 (checklist)

## Data Availability

The raw RNA sequencing data for the detection of NTRK fusions were deposited in the SRA/BioProject database (accession number: PRJNA715305; URL: http://www.ncbi.nlm.nih.gov/bioproject/715305). All other data are available from the corresponding authors (JHK or Y-LC) upon reasonable request.

## Patient consent for publication

Not required.

## Data availability statement

The raw RNA sequencing data for the detection of *NTRK* fusions were deposited in the SRA/BioProject database (accession number: PRJNA715305; URL: http://www.ncbi.nlm.nih.gov/bioproject/715305). All other data are available from the corresponding authors (JHK or Y-LC) upon reasonable request.

