## Supplementary Figures S1-S9 for "*NTRK* oncogenic fusions are exclusively associated with the serrated neoplasia pathway in the colorectum and begin to occur in sessile serrated lesions"

### Supplementary Figure S1. Sample collection workflow

(Abbreviations: CRC, colorectal cancer; SNUH, Seoul National University Hospital; MSI, microsatellite instability; MSS, microsatellite stable; MMR, mismatch repair; IHC, immunohistochemistry; FFPE, formalin-fixed paraffin-embedded; SSL, sessile serrated lesion; SSA, sessile serrated adenoma; SSP, sessile serrated polyp; SSLD, sessile serrated lesion with dysplasia; SSAD, sessile serrated adenoma with dysplasia; TSA, traditional serrated adenoma; HP, hyperplastic polyp; TALG, tubular adenoma with low-grade dysplasia; TVALG, tubulovillous adenoma with low-grade dysplasia; VALG, villous adenoma with low-grade dysplasia; TAHG, tubular adenoma with high-grade dysplasia; TVAHG, tubulovillous adenoma with high-grade dysplasia; VAHG, villous adenoma with high-grade dysplasia; IMC, intramucosal carcinoma; CALG, conventional adenoma with low-grade dysplasia; CAHG, conventional adenoma with high-grade dysplasia)

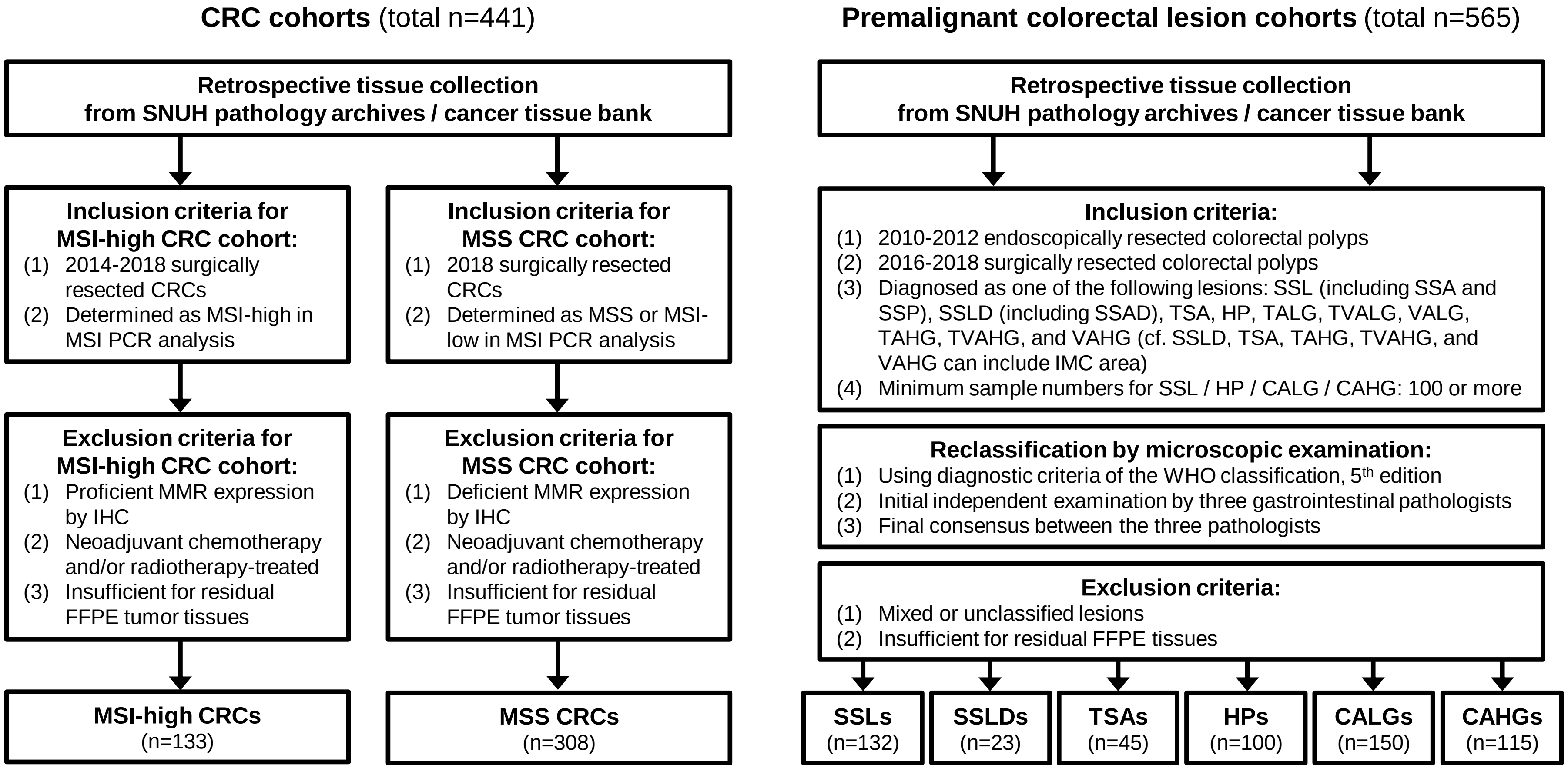

**Supplementary Figure S2. A novel *SFPQ-NTRK1* fusion identified in whole exome sequencing data**
Nineteen supporting reads mapped to the *SFPQ-NTRK1* fusion breakpoint identified in whole exome sequencing data using GeneFuse are shown. The base color of the read represents the quality score: green/blue, black, and yellow/red represent high, moderate, and low quality, respectively.

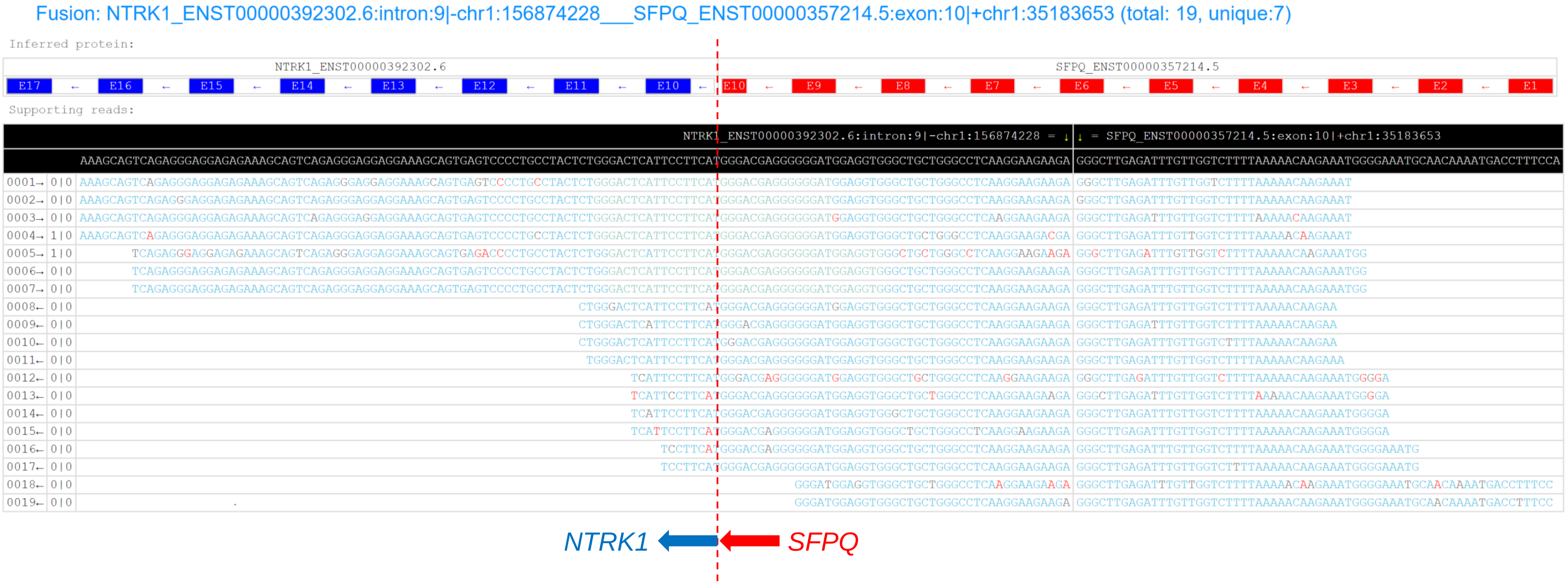

**Supplementary Figure S3. Photomicrographs of pan-TRK IHC in MSI-high CRCs harboring *NTRK* fusions (except for *ETV6-NTRK3*)**

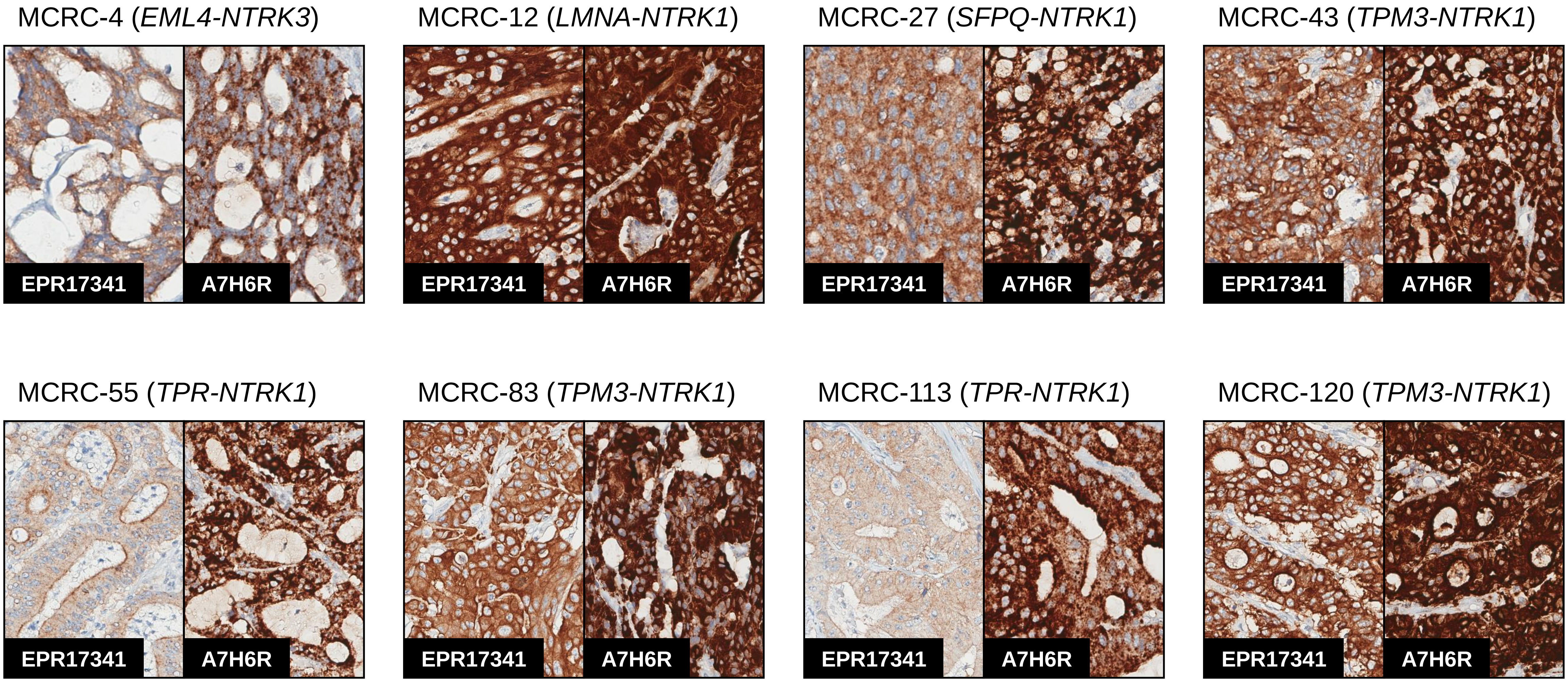

**Supplementary Figure S4. MLH1 proficiency in TRK-positive sessile serrated lesions with dysplasia (SSLDs)**
MLH1 IHC staining was positive in epithelial cell nuclei of the two TRK-positive SSLDs.

SSLD-10

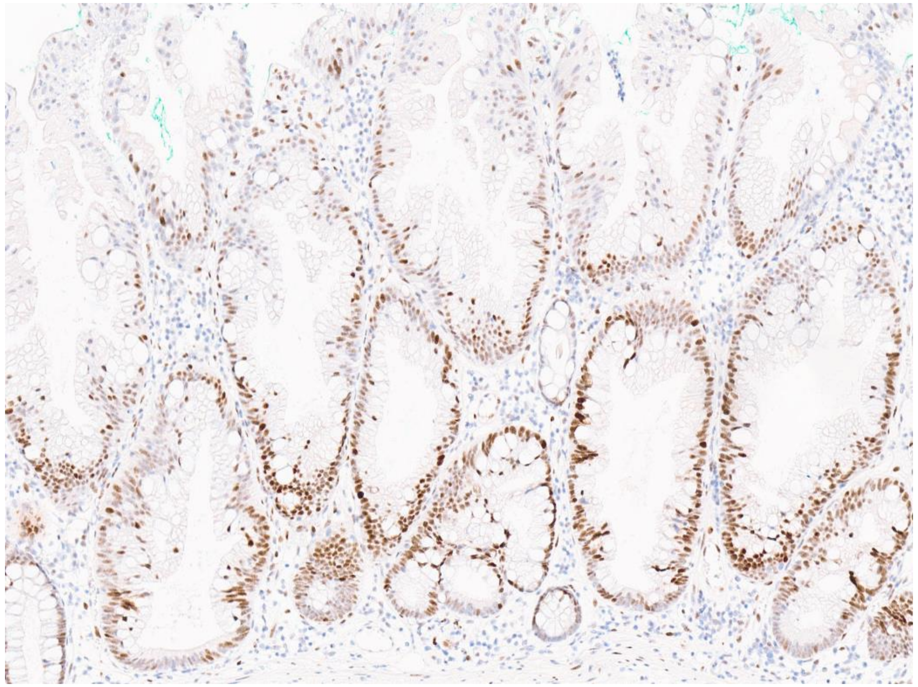

SSLD-13

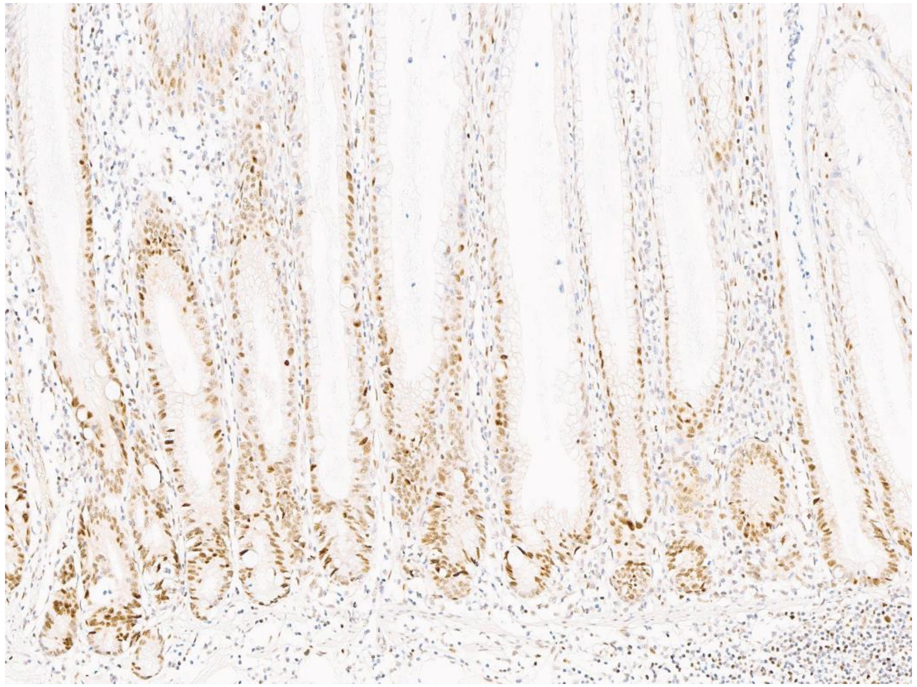

**Supplementary Figure S5. MLH1 proficiency in TRK-positive sessile serrated lesions (SSLs)**
MLH1 IHC staining was positive in epithelial cell nuclei of the four TRK-positive SSLs.

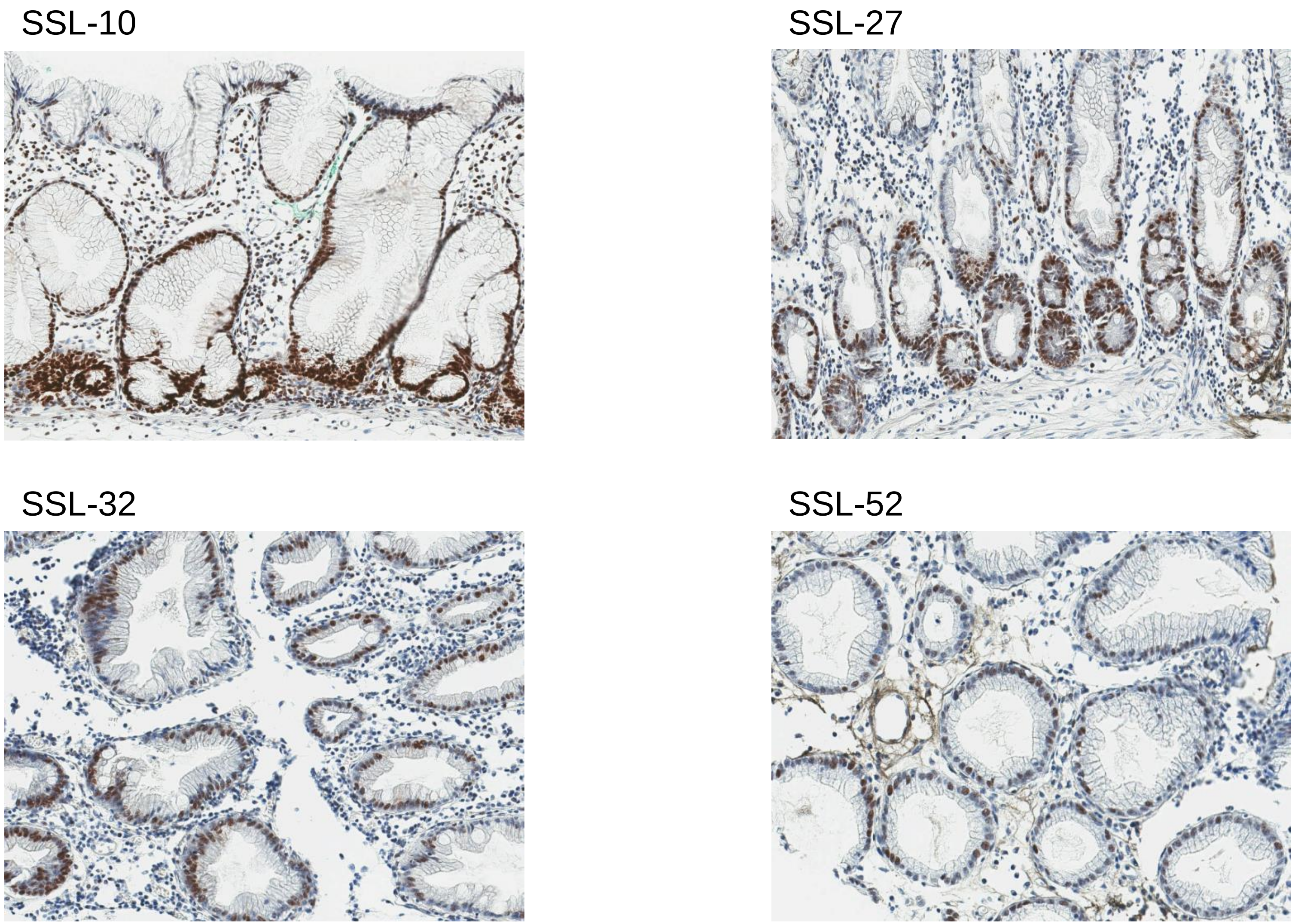

**Supplementary Figure S6. Histologic feature supporting the 'tracing serrated neoplastic crypts' hypothesis**
There is a serrated crypt at the lesion border area of a TRK-positive SSL, which shows focal weak staining of pan-TRK IHC.

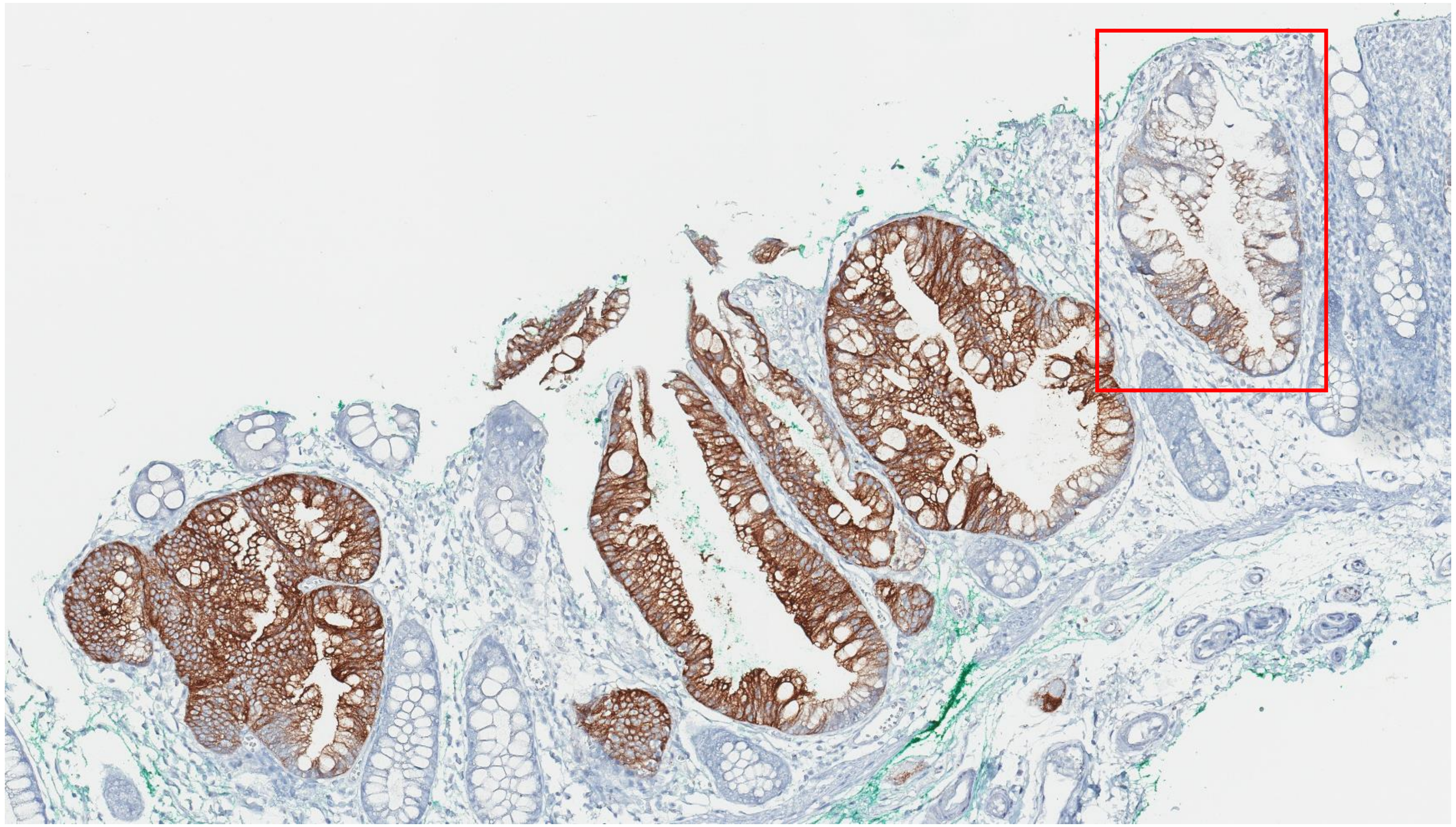

**Supplementary Figure S7. Histologic feature supporting the ‘causing serrated neoplastic crypts’ hypothesis**
In a TRK-positive SSL, there is a transitional morphological (mixed serrated and non-serrated morphological) crypt showing selective staining of pan-TRK IHC at the serrated morphologic area within the cross-sectioned crypt.

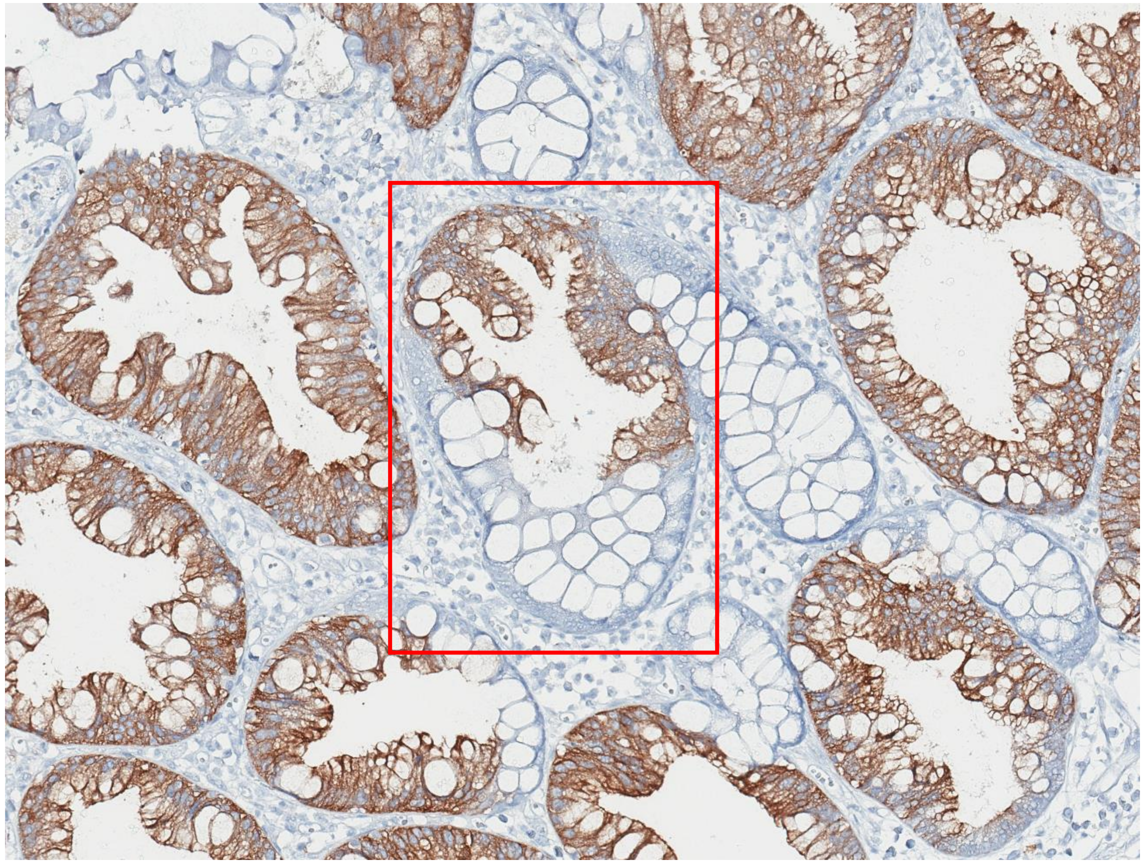

**Supplementary Figure S8. An ALK-positive sessile serrated lesion (SSL)**
Among the 132 SSLs tested for ALK immunohistochemistry using a companion diagnostic antibody (Ventana D5F3 CDx Assay, Roche, Basel, Switzerland), one case showed ALK positive staining in the epithelial cells, suggesting the presence of an *ALK* oncogenic fusion in the SSL.

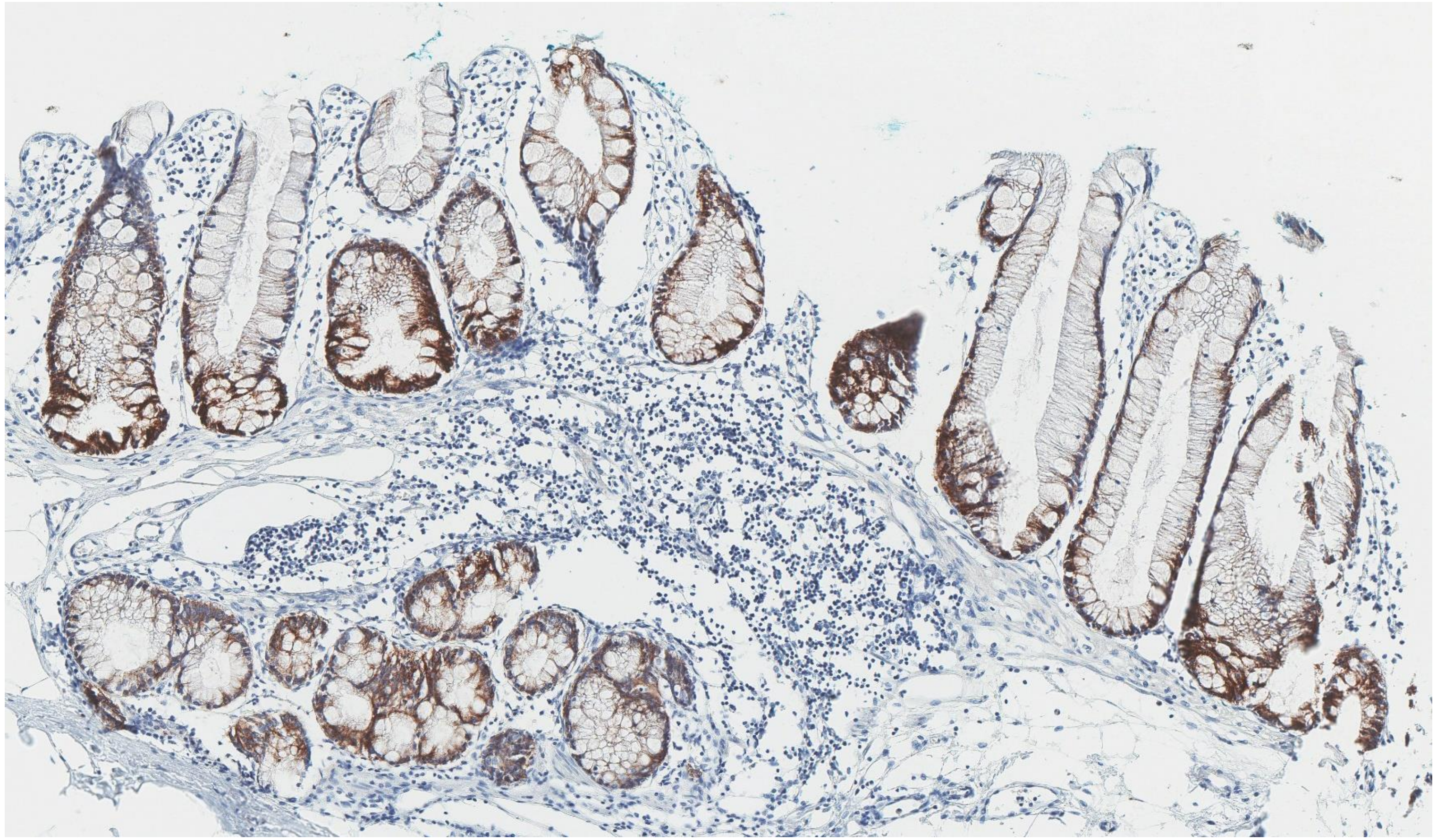

**Supplementary Figure S9. Partial loss of MLH1 expression in a sessile serrated lesion (SSL) with *MLH1* promoter methylation**
Note that the loss of MLH1 IHC expression was localized in the focal crypt base area within an *MLH1*-methylated SSL.

(Under the permission of the Journal of Pathology and Translational Medicine (JPTM), these figures were reused from Supplementary Figures previously used in our original article published in JPTM (Lee JA, Park HE, Yoo SY, et al. CpG island methylation in sessile serrated adenoma/polyp of the colorectum: implications for differential diagnosis of molecularly high-risk lesions among non-dysplastic sessile serrated adenomas/polyps. J Pathol Transl Med 2019;53: 225-35).)

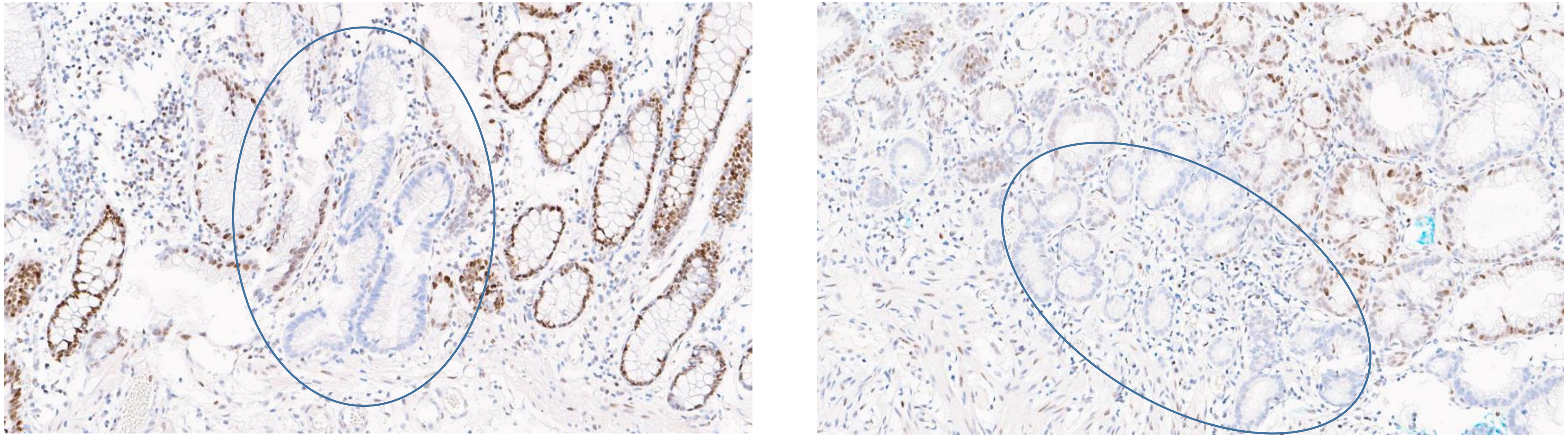
