## Supplementary Tables S1-S2 for "*NTRK* oncogenic fusions are exclusively associated with the serrated neoplasia pathway in the colorectum and begin to occur in sessile serrated lesions"

**Supplementary Table S1.** Comparison of clinicopathological, molecular, and immunological features between *NTRK* fusion-positive and -negative sporadic (*MLH1*-methylated) MSI-high CRCs (n = 58)

| Variable | | *NTRK* fusion-positive sporadic MSI-high CRCs (n = 11) | *NTRK* fusion-negative sporadic MSI-high CRCs (n = 47) | *P*-value |
|---|---|---|---|---|
| Age[a] | Older (≥ 72 years) | 8 (73%) | 26 (55%) | 0.333 |
| | Younger (< 72 years) | 3 (27%) | 21 (45%) | |
| Sex | Male | 3 (27%) | 11 (23%) | 1 |
| | Female | 8 (73%) | 36 (77%) | |
| Tumor location | Right-sided colon | 11 (100%) | 43 (91%) | 1 |
| | Left-sided colon or rectum | 0 (0%) | 4 (9%) | |
| Gross tumor type | Polypoid or fungating | 5 (45%) | 31 (66%) | 0.302 |
| | Ulceroinfiltrative | 6 (55%) | 16 (34%) | |
| Tumor size[b] | Larger (≥ 6.4 cm) | 6 (55%) | 19 (40%) | 0.504 |
| | Smaller (< 6.4 cm) | 5 (45%) | 28 (60%) | |
| Depth of invasion (pT) | Within the proper muscle (pT1/pT2) | 1 (9%) | 9 (19%) | 0.669 |
| | Beyond the proper muscle (pT3/pT4) | 10 (91%) | 38 (81%) | |
| Lymph node metastasis (pN) | Absent (pN0) | 8 (73%) | 37 (79%) | 0.696 |
| | Present (pN1/pN2) | 3 (27%) | 10 (21%) | |
| Distant metastasis (cM or pM) | Absent | 11 (100%) | 45 (96%) | 1 |
| | Present | 0 (0%) | 2 (4%) | |
| Lymphatic invasion | Absent | 5 (45%) | 35 (74%) | 0.079 |
| | Present | 6 (55%) | 12 (26%) | |
| Venous invasion | Absent | 11 (100%) | 44 (94%) | 1 |
| | Present | 0 (0%) | 3 (6%) | |
| Perineural invasion | Absent | 8 (73%) | 40 (85%) | 0.381 |
| | Present | 3 (27%) | 7 (15%) | |
| Tumor differentiation | Low grade (well or moderately differentiated) | 5 (45%) | 32 (68%) | 0.181 |
| | High grade (poorly differentiated) | 6 (55%) | 15 (32%) | |
| Mucinous histology | Non-mucinous carcinoma | 10 (91%) | 35 (74%) | 0.426 |

| | | | | |
|---|---|---|---|---|
| | Mucinous carcinoma | 1 (9%) | 12 (26%) | |
| Signet ring cell component | Absent | 11 (100%) | 41 (87%) | 0.583 |
| | Present | 0 (0%) | 6 (13%) | |
| Medullary component | Absent | 8 (73%) | 35 (74%) | 1 |
| | Present | 3 (27%) | 12 (26%) | |
| Tumor budding | Low or intermediate | 7 (64%) | 36 (77%) | 0.45 |
| | High | 4 (36%) | 11 (23%) | |
| Poorly differentiated clusters | Grade 1 or 2 | 5 (45%) | 32 (68%) | 0.181 |
| | Grade 3 | 6 (55%) | 15 (32%) | |
| Desmoplastic reaction | Mature or intermediate | 10 (91%) | 45 (96%) | 0.474 |
| | Immature | 1 (9%) | 2 (4%) | |
| *KRAS* or *BRAF* mutation | Absent | 11 (100%) | 27 (57%) | 0.011 |
| | Present | 0 (0%) | 20 (43%) | |
| CIMP | CIMP-high | 11 (100%) | 33 (70%) | 0.05 |
| | CIMP-low/negative | 0 (0%) | 14 (30%) | |
| *MLH1* promoter methylation | Methylated | 11 (100%) | 47 (100%) | NA |
| | Unmethylated | 0 (0%) | 0 (0%) | |
| *CACNA1G* promoter methylation | Methylated | 11 (100%) | 31 (66%) | 0.025 |
| | Unmethylated | 0 (0%) | 16 (34%) | |
| *SOCS1* promoter methylation | Methylated | 3 (27%) | 20 (43%) | 0.499 |
| | Unmethylated | 8 (73%) | 27 (57%) | |
| *CRABP1* promoter methylation | Methylated | 11 (100%) | 38 (81%) | 0.184 |
| | Unmethylated | 0 (0%) | 9 (19%) | |
| *RUNX3* promoter methylation | Methylated | 8 (73%) | 24 (51%) | 0.314 |
| | Unmethylated | 3 (27%) | 23 (49%) | |
| *IGF2* promoter methylation | Methylated | 10 (91%) | 29 (62%) | 0.082 |
| | Unmethylated | 1 (9%) | 18 (38%) | |
| *CDKN2A* promoter methylation | Methylated | 9 (82%) | 28 (60%) | 0.296 |
| | Unmethylated | 2 (18%) | 19 (40%) | |
| *NEUROG1* promoter methylation | Methylated | 11 (100%) | 34 (72%) | 0.055 |

| | | | | |
|---|---|---|---|---|
| | Unmethylated | 0 (0%) | 13 (28%) | |
| $CD3^+$ TIL density[c] | High (≥ 491 cells/mm$^2$) | 4 (40%) | 16 (35%) | 0.733 |
| | Low (< 491 cells/mm$^2$) | 6 (60%) | 30 (65%) | |
| $CD8^+$ TIL density[c] | High (≥ 276 cells/mm$^2$) | 4 (40%) | 13 (28%) | 0.471 |
| | Low (< 276 cells/mm$^2$) | 6 (60%) | 33 (72%) | |
| $FoxP3^+$ TIL density[c] | High (≥ 107 cells/mm$^2$) | 3 (27%) | 14 (30%) | 1 |
| | Low (< 107 cells/mm$^2$) | 8 (73%) | 33 (70%) | |
| $CD68^+$ TAM density[c] | High (≥ 1019 cells/mm$^2$) | 6 (55%) | 21 (45%) | 0.555 |
| | Low (< 1019 cells/mm$^2$) | 5 (45%) | 26 (55%) | |
| $CD163^+$ TAM density[c] | High (≥ 627 cells/mm$^2$) | 6 (55%) | 21 (45%) | 0.555 |
| | Low (< 627 cells/mm$^2$) | 5 (45%) | 26 (55%) | |
| TLS activity[d] | Active (maximum diameter of LAs ≥ 1 mm) | 6 (55%) | 23 (49%) | 0.738 |
| | Inactive (maximum diameter of LAs < 1 mm) | 5 (45%) | 24 (51%) | |
| PD-L1 expression in tumor cells | Low (H-score < 50) | 9 (82%) | 38 (81%) | 1 |
| | High (H-score ≥ 50) | 2 (18%) | 9 (19%) | |
| PD-L1 expression in immune cells | Low (H-score < 100) | 5 (45%) | 24 (51%) | 0.738 |
| | High (H-score ≥ 100) | 6 (55%) | 23 (49%) | |

Abbreviations: MSI-high, microsatellite instability-high; CRC, colorectal cancer; CIMP, CpG island methylator phenotype; NA, not applicable; TIL, tumor-infiltrating lymphocyte; TAM, tumor-associated macrophage; TLS, tertiary lymphoid structure; LAs, lymphoid aggregates.

[a]Age subgrouping was performed using an average age of the 58 MSI-high CRCs.

[b]Tumor size subgrouping was performed using an average value of maximum diameters of the 58 MSI-high CRCs.

[c]Density subgrouping of each TIL or TAM was performed using an average density of each TIL or TAM in the 58 MSI-high CRCs.

[d]TLS activity of each MSI-high CRC was classified into active or inactive according to the Ueno criteria (Ueno H et al. Am J Clin Pathol. 2013 Apr;139(4):434-41. doi: 10.1309/AJCPWHUEFTGBWKE4).

**Supplementary Table S2.** Comparison of clinicopathological and molecular features between *NTRK* rearrangement-positive and -negative SSLs (n = 132)

| Variable | | *NTRK* rearrangement-positive SSLs (n = 5) | *NTRK* rearrangement-negative SSLs (n = 127) | *P*-value |
|---|---|---|---|---|
| Age | Older (≥ 50 years) | 5 (100%) | 106 (83%) | 1 |
| | Younger (< 50 years) | 0 (0%) | 21 (17%) | |
| Sex | Male | 3 (60%) | 83 (65%) | 1 |
| | Female | 2 (40%) | 44 (35%) | |
| Lesion location | Right-sided colon | 5 (100%) | 102 (80%) | 0.583 |
| | Left-sided colon or rectum | 0 (0%) | 25 (20%) | |
| Lesion size | Larger (≥ 10 mm) | 5 (100%) | 50 (39%) | 0.011 |
| | Smaller (< 10 mm) | 0 (0%) | 77 (61%) | |
| Synchronous multiplicity of SLs | Single SSL only | 3 (60%) | 68 (54%) | 1 |
| | Multiple SLs | 2 (40%) | 59 (46%) | |
| *KRAS* or *BRAF* mutation[a] | Absent | 5 (100%) | 71 (66%) | 0.17 |
| | Present | 0 (0%) | 37 (34%) | |
| CIMP | CIMP-high | 4 (80%) | 29 (23%) | 0.014 |
| | CIMP-low/negative | 1 (20%) | 98 (77%) | |
| *MLH1* promoter methylation | Methylated | 1 (20%) | 6 (5%) | 0.242 |
| | Unmethylated | 4 (80%) | 121 (95%) | |
| *CACNA1G* promoter methylation | Methylated | 4 (80%) | 48 (38%) | 0.078 |
| | Unmethylated | 1 (20%) | 79 (62%) | |
| *SOCS1* promoter methylation | Methylated | 2 (40%) | 17 (13%) | 0.151 |
| | Unmethylated | 3 (60%) | 110 (87%) | |
| *CRABP1* promoter methylation | Methylated | 4 (80%) | 77 (61%) | 0.648 |
| | Unmethylated | 1 (20%) | 50 (39%) | |
| *RUNX3* promoter methylation | Methylated | 4 (80%) | 32 (25%) | 0.02 |
| | Unmethylated | 1 (20%) | 95 (75%) | |
| *IGF2* promoter methylation | Methylated | 3 (60%) | 38 (30%) | 0.173 |
| | Unmethylated | 2 (40%) | 89 (70%) | |

| | | | | |
|---|---|---|---|---|
| *CDKN2A* promoter methylation | Methylated | 5 (100%) | 50 (39%) | 0.011 |
| | Unmethylated | 0 (0%) | 77 (61%) | |
| *NEUROG1* promoter methylation | Methylated | 5 (100%) | 73 (57%) | 0.078 |
| | Unmethylated | 0 (0%) | 54 (43%) | |

Abbreviations: SSLs, sessile serrated lesions; SLs, serrated lesions (including hyperplastic polyp, traditional serrated adenoma, and sessile serrated lesion); CIMP, CpG island methylator phenotype.
[a]19 cases were excluded in *KRAS* or *BRAF* mutation testing due to inadequate quality or quantity of their extracted DNA samples.
